# The genetic relationships between post-traumatic stress disorder and its corresponding neural circuit structures

**DOI:** 10.1101/2024.08.25.24312540

**Authors:** Qian Gong, Honggang Lyu, Lijun Kang, Simeng Ma, Nan Zhang, Xin-hui Xie, Enqi Zhou, Zipeng Deng, Jiewei Liu, Zhongchun Liu

**Author notes:** Contributing authors.

## Abstract

Post-traumatic stress disorder (PTSD) may be linked to abnormalities in neural circuits that facilitate fear learning and memory processes. The precise degree to which this connection is influenced by genetic factors is still uncertain. This study aimed to investigate the genetic association between PTSD and its corresponding brain circuitry components. We first conducted a meta-analysis using the summary of PTSD genome-wide association studies (GWAS) from multiple cohorts to enhance statistical power (sample size = 306,400). Then, based on the result of the GWAS meta-analysis, and utilizing the lifetime trauma events (LTE) trait as a control for PTSD, we proceeded with subsequent investigations. We investigated the genetic association of PTSD and LTE with nine brain structure traits related to the brain circuitry by various methodologies, including heritability tissue enrichment analysis, global and local genetic correlations, polygenic overlap analysis, and causal inference. As a result, we discovered an enrichment of heritability for PTSD within circuitry-relevant brain regions such as the cingulate cortex and frontal cortex, alongside the identification of weak genetic correlations between PTSD and these brain regions. We have observed a polygenic overlap between the two trauma-related traits and nine traits of brain circuitry components such as global cortical area and cingulum. A total of 31 novel jointly significant genetic loci (conjunction FDR ***<*** 0.05) associated with PTSD and nine brain structures were identified, suggesting a potential connection between them, and these loci are involved in the process of DNA damage and repair as well as the pathway of neurodegenerative diseases. We also identified a potential causal relationship between PTSD and the surface area of the frontal pole. Our findings offer a valuable understanding of the genetic mechanisms underlying PTSD and its associated brain circuitry.

## 1 Introduction

Post-traumatic stress disorder (PTSD) is a severe mental disorder that may occur after exposure to traumatic life events. It is characterized by intrusive symptoms, avoidance of trauma-related cues, hyperarousal, negative cognition and moods as the core symptoms.[1] The prevalence of PTSD in the general population is approximately 6%. However, people who have experienced various types and degrees of trauma may exhibit higher prevalence rates. For example, war veterans and victims of assault may experience a prevalence of 25% to 35%.[2–4] It has led to serious social burdens, including hospitalizations, suicides and substance abuse.[5] Identifying the mechanisms linked to PTSD would enable the development of interventions to safeguard vulnerable groups and address the disorder, thereby alleviating the economic and emotional strain. Prior studies have indicated that the development of PTSD may be associated with abnormalities in brain circuitry that facilitate fear learning and memory mechanisms.[6, 7] The circuit is mainly composed of the amygdala, hippocampus, prefrontal cortex (PFC) and the connections between them.[6, 7] Researchers suggest that (1) hyperresponsiveness in amygdala is associated with promoting fear associations and expression of fear responses; (2) defects in hippocampal function are associated with mediating deficits in appreciation of safe contexts and explicit learning and memory; and (3) defects in frontal cortex function are associated with mediating deficits in extinction and the capacity to suppress attention or response to trauma-related stimuli.[8, 9]

Both animal experiments and neuroimaging studies have validated these views that PTSD is associated with abnormalities in the neural circuits.[8, 10–12] In the neural circuit, amygdala plays a role in fear processing. It was found in rodent studies that sensory information goes first to the basolateral amygdala, where fear learning occurs, and then the signal is transmitted to the central amygdala, which regulates the expression of fear-related behaviors.[13, 14] And in neuroimaging studies, individuals with PTSD exhibit heightened activation in the amygdala, which might lead to an overreaction to fear.[15, 16] Hippocampus plays a role in fear acquisition, memory retention, and expression processes, stimulation of which can alter the recall of fear extinction memory.[17, 18] Researchers have identified reduced hippocampal neurogenesis and dendritic spine loss in the hippocampal CA3 region in trauma-model rodent studies.[19] Meanwhile, neuroimaging studies demonstrate a similar reduction in hippocampal volume among individuals diagnosed with PTSD.[20, 21] PFC plays a role in decision-making and executive functions. Its subregions are associated with suppressing the expression of fear behaviors and the retention of fear extinction memories.[22, 23] This is consistent with reduced gray matter volume and reduced function in the PTSD neuroimaging studies.[24–27] Moreover, the bidirectional communication of the PFC with the hippocampus and amygdala is also important to regulate traumatic fear learning and its extinction.[28] The structural connections between amygdala, hippocampus, and prefrontal structure, specially the microstructure of the cingulum bundle (CG), fornix/stria terminalis (FST), and uncinate fasciculus (UNC), also show abnormalities in patients with PTSD.[29, 30]

The advent of large-scale magnetic resonance imaging (MRI) and genetic datasets has enabled researchers to gain insight into alterations in brain structure and the nexus between disorders and genetic underpinnings.[31] Several twin studies indicate a moderate heritability of brain structure.[32–34] Genome-wide association studies (GWAS) serve as valuable tools for elucidating the genetic mechanisms underlying imaging alterations from a genetic vantage point. Certain single nucleotide polymorphisms (SNPs) identified in these studies delineate factors associated with clinical illness such as schizophrenia, while others delineate risk or protective elements for psychiatric disorders such as neurodevelopment.[35–38]. Consequently, a growing number of studies have begun to focus on the genetic associations and polygenic links between psychiatric disorders and structural brain phenotypes. Most of these studies hypothesized that pleiotropic genes encode risk for mental illness through effects on intermediate or endophenotypes of brain structure.[39–42] This is helpful to quantify the weight of brain traits in the psychiatry disease, discover new loci to complement previous results, and explore the mechanisms by which brain phenotypes act as intermediate or endophenotypes through the biological functions of shared loci.[39–42]

There has been a lack of exploration between PTSD and brain structural studies. By integrating advanced statistical models and the results of large-scale GWAS meta-analyses, it is possible to investigate the genetic overlap between PTSD and brain imaging traits, thereby enhancing the capability to identify associated loci. It is meaningful to study the genetic relationship between PTSD and the neural circuits comprising the amygdala, hippocampus, and PFC and their connections, considering the consistent correlation in clinical symptoms, rodent studies, and neuroimaging studies.[43] Moreover, in GWAS studies of PTSD, the loci identified were also associated with clinical manifestations or risk factors for psychiatric disorders[44, 45], which is consistent with GWAS studies of brain traits as mentioned before. Therefore, we consider that the study of the co-inheritance or polygenicity of PTSD and brain structure traits related to the neurocircuitry can facilitate the identification of new loci and biological mechanisms of PTSD initiation through structural changes in the brain.

In this work, we first combined the GWAS summary data for PTSD from different organizations. We also included lifetime trauma events (LTE) as a control, considering that traumatic events may directly cause changes in brain structure rather than being linked to the development of PTSD. Second, We combined the two trauma-related GWAS summaries with brain cell-type-specific gene expression data to determine if the heritability of trauma-related phenotypes enriches the brain circuits described above. Third, we added GWAS data of brain structure traits (see Methods for details) to examine the genetic correlations and overlap between trauma-related and them. We use MiXeR[46] to determine the degree of overlapping genetic architecture. We identified risk loci shared between them using the conjunctional false discovery rate (conjFDR)[47] and applied Functional Mapping and Annotation (FUMA) to annotate the identified loci to determine molecular functions of shared risk variants for trauma and brain structure traits.[48] Last, we used bidirectional two-sample Mendelian randomization to investigate the causal relationship between these two types of traits.

## 2 Methods and Materials

### 2.1 GWAS Data and Meta-analysis

GWAS summary data for PTSD was obtained from three large studies: (1) a meta-analysis combining 51 cohorts of European ancestry, including the Psychiatric Genomics Consortium (PGC, freeze2 PGC-PTSD meta-analysis), and the UK Biobank, encompassing a total of 182,199 samples.[44] (2) Million Veteran Program (MVP, 36,301 cases, 178,107 controls)[49] and (3) FinnGen (2,282 cases, 337,577 controls); we utilized liftOver to convert the summary data from the hg38 build to hg37. In the first meta-analysis of 51 cohorts, 19 methods were used to study PTSD, the most common being the Clinician-Administered PTSD Scale and PTSD Checklist, and 91% of participants with PTSD phenotypes were using the PTSD symptom score, while the others used the case/control status.[44] Specific methods of participant and phenotype selection can be found in the original article.[44, 45] PTSD in MVP was a diagnosis of case/control applying the algorithm based on the electronic medical record.[50] PTSD in FinnGen was similarly defined as a case/control status. The metaanalysis of these three summaries was performed using METAL.[51]. We combined p-values across studies taking into account a study specific weight and direction of effect. Risk loci were defined by FUMA[48], using the default parameters.

Considering that changes in brain structure may be directly caused by trauma and that there are many people who experience trauma events and do not develop PTSD, we used the GWAS summary for LTE from the UK biobank as a reference.[44] The researchers constructed a count measure of LTE from 8 trauma items of the selfreported retrospective trauma screener from the UKBB mental-health questionnaire.

GWAS summary data for cortical was obtained from ENIGMA3 (https://enigma.ini.usc.edu/). It comprised results from 33,992 participants of European ancestry (23,909 from 49 cohorts in the ENIGMA consortium and 10,083 from the UK Biobank).[35] The traits of this study involve the surface area (SA) and thickness (TH) of 34 brain regions of the Desikan-Killiany (DK) atlas. These brain regions were divided according to the gyral-based neuroanatomical regions.[52] According to the radial unit hypothesis, neural progenitor cell proliferation leads to the increase in cortical SA, while their neurogenic divisions influence TH.[53] The PFC was not clearly delineated on this atlas, and given the location of the prefrontal lobes and the regions of the Brodmann area (BA) atlas [54] that we would use in tissue enrichment, we used the frontal poles SA (FSA) and TH (FTH) instead of PFC. In addition, we also considered the relationship between global cortical SA (GSA) and thickness (GTH) and trauma traits.

We chose the volume of amygdala (AMY) and hippocampus (HIP) as the subcortical traits. GWAS for the volume of amygdala was from a formal study including 53 study samples from the CHARGE consortium, ENIGMA consortium, and UK Biobank.[36] The traits were defined as the mean volume (in *cm*^3^) of the left and right amygdala.[36] Segmentation of brain regions was done according to the freely available and in-house segmentation methods that come with the software used for the different cohorts.[36] GWAS for the volume of hippocampus was obtained from a GWAS study of 33,536 individuals in the ENIGMA Consortium and the CHARGE Consortium.[37] Hippocampal volumes were also estimated using the automated segmentation algorithm in FMRIB Software Library (FSL) and FreeSurfer.[55, 56]

GWAS summary data for white matter was obtained from a GWAS of dMRI data from 43,802 individuals across five data resources.[38] This article included 5 DTI indicators of 21 white matter pathways. We selected CG, UNC, and FST which are related to PTSD. And we chose fractional anisotropy (FA) as the primary DTI indicator because it has received the most attention in brain white matter research.[57] dMRI measures the directional diffusion of water molecules in tissues, and FA quantifies the degree of directionality, with high FA indicating more organized structures.[57].

The specific locations of the above-mentioned brain areas are shown in Fig 1.

**Fig. 1.**
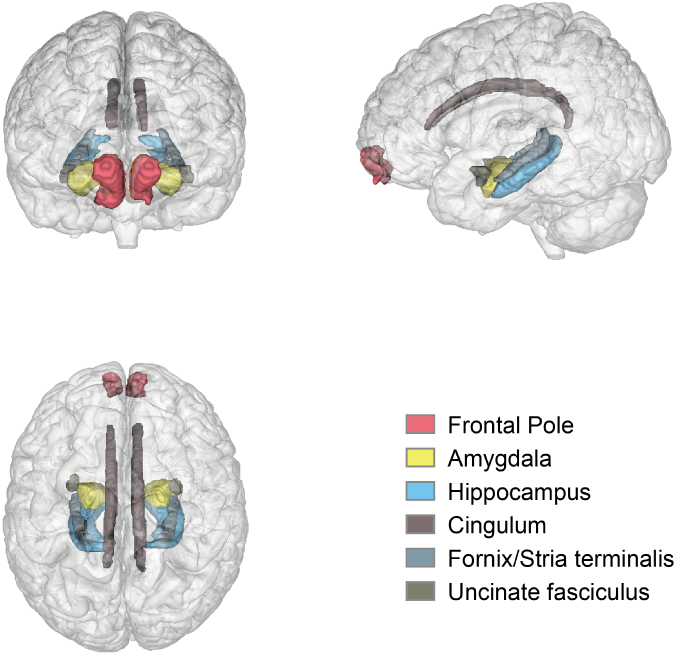
Brain regions associated with trauma neural circuits mentioned in this article. These brain regions encompass cortical, subcortical structures (amygdala and hippocampus), as well as white matter tracts (cingulum, fornix/stria terminalis, and uncinate fasciculus).

### 2.2 Statistical Analysis

#### 2.2.1 Brain tissue enrichment

We used linkage-disequilibrium (LD) score regression (LDSC) applied to specifically expressed genes (LDSC-SEG)[58], which tests for enrichment for per-SNP heritability to identify tissue-type-specific enrichment of SNPs. The 13 pre-computed cell-type of brain annotations in GTEx v8 were used in the analysis.[59] A Benjamini-Hochberg false discovery rate (BH-FDR) corrected *P* value of 0.05 was used as a significant threshold.

#### 2.2.2 Global correlations

Global correlations were conducted by LDSC[60]. SNPs with high LD exhibit higher *χ*^2^ statistics for polygenic traits compared to SNPs with low LD on average, and a similar pattern arises when replacing the single study statistic with the product of z-scores from two studies with a correlation. We used the calculated LD score from the 1000 Gene European population[61] to calculate the global correlation (*r_g_*) between PTSD and brain structure traits. Only SNPs in HAPMAP3 (MAF *<* 0.01 and INFO *>* 0.9) were used in the calculation. The range of *r_g_* is between −1 and 1, with −1 indicating a perfect negative correlation and 1 indicating a perfect positive correlation. The BH-FDR corrected *P* value of 0.05 was used as a significant threshold (FDR *P <* 0.05).

#### 2.2.3 Local correlations

Local genetic correlations were calculated by Local Analysis of [co]Variant Association (LAVA).[62] Traditional genetic correlation analysis uses a genome-wide average approach, which may overlook localized genetic contributions and hinder the detection of correlations driven by opposing or heterogeneous effects across the genome. LAVA can estimate genetic relatedness within specific regions of the genome, allowing for inverse correlations between different loci. For LAVA analysis, we followed the protocol described in the original article using the LD reference panel based on 1000 Genomes phase 3 genotype data for European samples,[61, 62] and the partition of the genome into 2495 regions with an average size of 1 Mb. Only regions revealing significant after Bonferroni corrected estimated SNP heritability (*P <* 0.05*/*2495) in both traits were used to estimate bivariate local genetic correlations between the traits. The BH-FDR corrected *P* value of 0.05 was used as a significant threshold.

#### 2.2.4 Polygenic overlap and shared loci

To examine the genetic overlap between pairs of trauma-related phenotypes and brain imaging phenotypes, we used MiXeR v1.3[46] to estimate the overall shared polygenic structure regardless of effect direction and coefficients. To assess polygenic overlap between two traits, MiXeR calculates the overall count of shared and trait-specific causal variants—variants with a non-zero additive genetic effect on a trait. We first performed univariate analysis to derive the number of loci affecting each shape (i.e., polygenic) and the average magnitude of the additive genetic association between these variants (i.e., discoverability). Then, a bivariate analysis model was used to estimate the total number of shared and phenotype-specific causal variants. The results of MiXeR are presented as a Venn diagram of shared and unique polygenic components across traits. MiXeR was implemented to calculate the Dice coefficient (i.e., the ratio of shared variants to the total number of variants). The model fit evaluated via the Akaike information criterion (AIC) was based on the maximum likelihood of GWAS z-scores and was illustrated with conditional Q-Q plots.

To improve the discovery of specific genetic variants shared between phenotypes, we applied the conditional FDR (condFDR) and conjunction FDR (conjFDR) statistical frameworks.[47] condFDR improves the statistical power of association analysis by merging overlapping SNP associations, rearranging association statistics in primary traits according to test statistics for secondary traits, and vice versa.[47, 63] The maximum value between the condFDR for both traits was defined as conjFDR, which provides a conservative estimate for the detection of shared genetic variants. We removed the major histocompatibility complex (MHC) and chromosome 8p23.1 genomic regions (hg19, chr6:25,119,106–33,854,733 and chr8:7,242,715–12,483,982) from condFDR analysis to exclude the potential impact of their complex regional LD patterns. A threshold of statistical significance for identifying shared genetic variants was set at conjFDR *<* 0.05. A locus that did not physically overlap with findings from the original GWAS and the National Human Genome Research Institute-European Bioinformatics Institute (NHGRI-EBI)[64] GWAS Catalog, was considered novel.

#### 2.2.5 Functional annotation

We apply the FUMA[48] protocol with default parameters to define the independent loci and lead SNPs (conjFDR *P <* 0.05). For all candidate SNPs, which have an LD (*r*^2^ *≥* 0.6) with at least one of the independent significant SNPs, we conducted functional annotation analysis with the Combined Annotation Dependent Depletion (CADD)[65], RegulomeDB[66], and chromatin states[67]. We performed three gene mapping strategies for candidate SNPs, including positional mapping, expression quantitative trait locus (eQTL) association, and chromatin interaction mapping. We used information from GTEx v8[68] and PsychENCODE[69] to find out the expression of genes we had found. Then, we evaluated the effects’ direction of the shared loci by comparing their z-scores and odds ratios. Using Gene2Func in FUMA, we annotated the set of prioritized genes from the first step with biological functions and mechanisms. Adjusted P-values with the BH-FDR for multiple test correction methods were computed.

#### 2.2.6 Causal relationship

To test the potential causal relationship between trauma-related traits and brain structure, we conducted a bidirectional two-sample Mendelian randomization analysis. We selected significant independent SNPs (a window size of 1Mb, *r*^2^ threshold of 0.001 and *P* value threshold of 5e-8) from the GWAS summary mentioned before as the instrument variants (IVs). We calculate the F-statistics to test the strength of instrument.[70] We used the NHGRI-EBI GWAS catalog[64] and the PhenoScanner V2 database[71] to remove SNPs with the confounders and outcome. If an instrument SNP was not available in the outcome GWAS summary, we used LDlinkR[72] to identify a proxy SNP in LD with the target SNP (*r*^2^ *>* 0.8). We reported the results for inverse-variance weighted (IVW),[73], weighted median, and MR-Egger regression.[74, 75] We executed an MR-Egger regression to examine the potential bias of directional pleiotropy.[75] MR-Pleiotropy Residual Sum and Outlier (MR-PRESSO) method[76] was applied to detect and correct for horizontal pleiotropy. Last, we performed a leave-one-out analysis where one SNP was removed at a time and IVW was conducted based on the remaining SNPs.

## 3 Results

### 3.1 GWAS meta-analysis for PTSD

To enhance statistical power, we conducted a meta-analysis utilizing three reports available from multiple cohorts (sample size = 306,400). We identified 16 genome-wide significant (*P <* 5 *×* 10*^−^*^8^) risk loci for PTSD (Table 1 and Fig 2). The LDSC and quantile-quantile (Q-Q) plots showed that most of the associations were attributed to polygenicity rather than confounding factors. (intercept, 1.012, s.e., 0.007) (Supplementary Fig. 1). Among these 16 loci, four were reported by the first GWAS summary about PGC and UKBB[44], and two loci were reported by the second summary about MVP.[49]

**Fig. 2.**
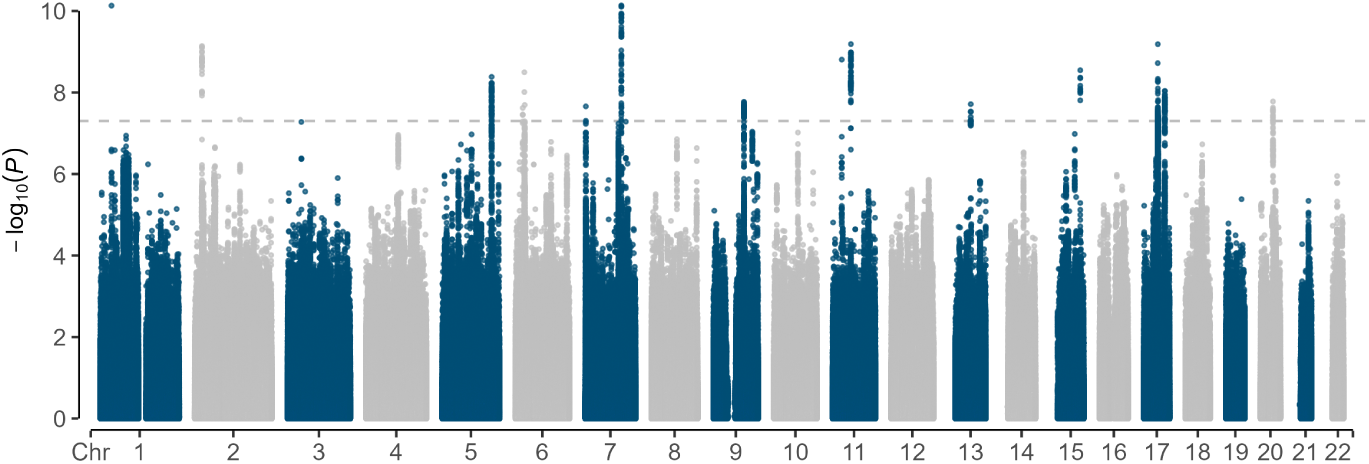
Manhattan plot for the GWAS meta-analysis. The associations are from the three GWAS meta-analysis. The gray line shows the genome-wide significant *P* threshold (*P <* 5 *×* 10*^−^*^8^).

**Table 1.**
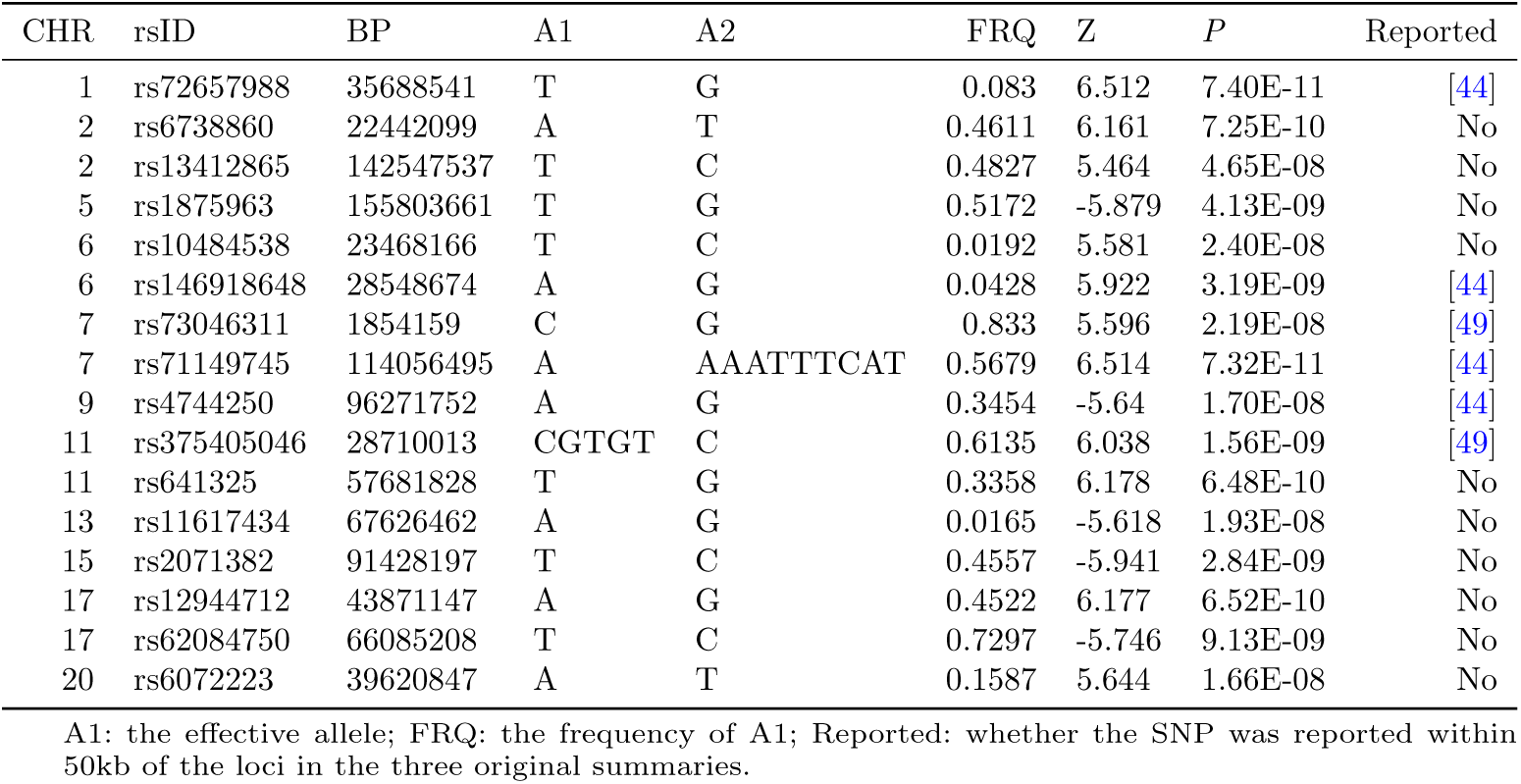
Genome-wide significant loci from PTSD GWASs meta analysis.

### 3.2 The tissue enrichment of heritability aligned with circuitry structure

To identify tissue types that are more likely to be involved in the neural circuit of trauma, we used the trauma-related GWAS summary statistics and 13 brain tissue type-specific data from GTEx v8 by LDSC-SEG. We found that the expression of PTSD loci was enriched in “Brain Anterior cingulate cortex (BA24)” (*P_F DR_* = 0.0012), “Brain Frontal Cortex (BA9)” (*P_F DR_* = 0.0019), and “Brain Cortex” (*P_F DR_* = 0.013), while the expression of LTE loci was enriched in “Brain Cortex” (*P* = 0.009, not significant after adjustment) (Supplementary Table 1). The results are consistent with the brain areas we mentioned earlier.

### 3.3 Global and local genetic correlations

We estimated the global genetic correlation using LDSC between trauma-related and brain structure-related traits. The results showed weak correlation between PTSD and GSA (*r_g_* = −0.0901, *P* = 0.018), LTE and FST (*r_g_* = −0.1301, *P* = 0.0035), LTE and CG (*r_g_* = −0.1038, *P* = 0.0224). The results were not significant after FDR (Supplementary Table 2).

Local genetic correlation may be overridden in the calculation of *r_g_*, so we used LAVA to estimate the local genetic correlations. LAVA local correlations provided further evidence of the relationship between trauma and brain structure. Loci with a significant amount of local genetic signal for both traits (*P <* 0.05/2495) were used in the bivariate analysis. PTSD is significantly correlated with the local genetic variations in GSA (n = 4), GTH (n = 1), FSA (n = 3), AMY (n = 3), HIP (n = 5), CG (n = 6), FST (n = 7), and UNC (n = 4). The number of loci demonstrating a significant local correlation between LTE and brain structure is fewer than those observed in PTSD. The results between LTE and brain structure were as follows: GTH (n = 1), CG (n = 2), and UNC (n = 2). Especially, the region in chr7: 155280611-156344386 is significant in both PTSD-GSA and PTSD-hippocampus pairs, and the region in chr17: 43460501-44865832 is significant in both PTSD-GSA and PTSD-FST. (Supplementary Table 3).

### 3.4 Shared genomic architectures and loci between trauma-related traits and brain structure-related traits

Univariate MiXeR estimated the SNP heritability (*h*^2^) to be 0.049 for PTSD and 0.066 for LTE. The *h*^2^ of brain structure estimated by MiXeR ranged from 0.0041 to 0.32. The results of trait-influencing variants showed that trauma-related traits were 3-4 times more polygenic than brain structure, with 10K and 7.8K variants relative to PTSD and LTE, while there were only 3.2K variants related to GTH, which was the most polygenic brain structure-related phenotype. (Supplementary Fig. 2 and Supplementary Table 4). Bivariate MiXeR revealed significant polygenic overlap between brain structure-related variants and trauma-related variants. The UNC shared the largest proportion of trait-influencing variants with PTSD (98%, 2008 out of 2059), and the GSA shared the largest proportion of trait-influencing variants with LTE (96%, 1751 out of 1820). (Fig 3) Among the pairs of PTSD, the results of GSA, GTH, CG, FST, and UNC can also be further supported by AIC values and Q-Q diagrams (Supplementary Fig. 3). All the results can be found in Supplementary Table 5.

**Fig. 3.**
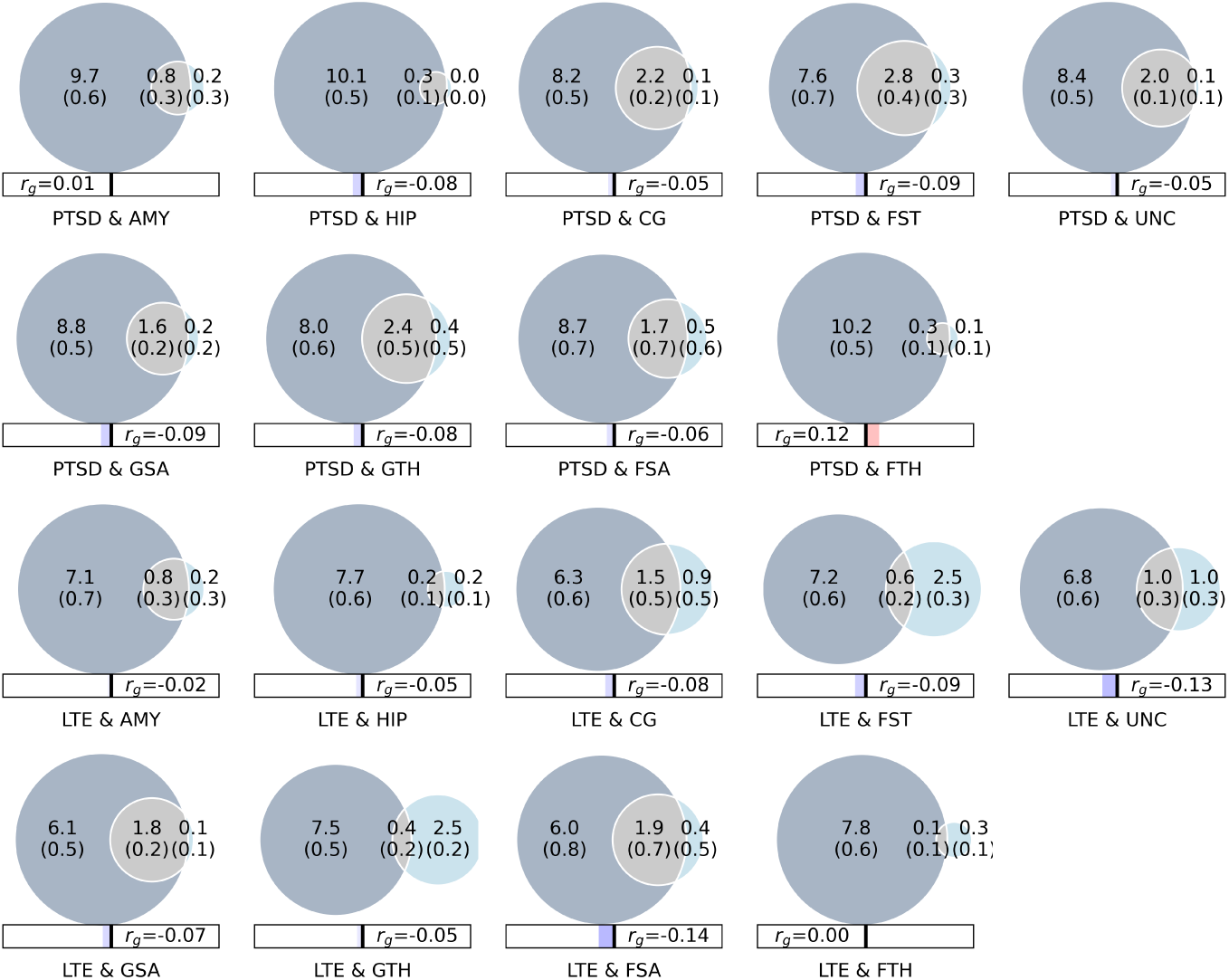
Polygenic overlap between trauma-related traits and brain structure-related traits. The Venn diagram shows the estimated number of causal variants shared between trauma-related traits and brain structure-related traits. The number of causal variants in thousands is shown, as well as the standard error. Abbreviations: PTSD, post-traumatic stress disorder; LTE, lifetime trauma events; GSA, global cortical surface area; GTH, global cortical thickness; FSA, frontal poles surface area; FTH, frontal poles thickness; AMY: amygdala; HIP: hippocampus; CG: cingulum bundle; FST: fornix/stria terminalis; UNC: uncinate fasciculus.

To explore the genetic sharing of trauma-related traits and brain structure-related traits, we applied conjFDR analysis for each pair. We discovered 40 independent loci that were jointly associated with PTSD with conjFDR *<* 0.05, including 12 loci for GSA, 7 loci for GTH, 1 locus for FSA, 2 loci for FTH, 1 locus for AMY, 4 loci for HIP, 11 loci for CG, 1 locus for FST, and 2 loci for UNC (Fig 4 and Table 2). In particular, one locus (lead SNP rs2352974, chr3:49734229-50176259) was associated in both CG and UNC with PTSD. There were only 9 loci that were jointly associated with LTE with conjFDR *<* 0.05. Therefore, in the subsequent functional analysis, we will only show PTSD-related results. Among those 40 PTSD risk loci identified by leveraging GWAS of brain structural traits, 31 lead SNPs that had not been reported as significant in the previous PTSD GWAS studies were defined as novel. See Supplementary Table 6 for details.

**Fig. 4.**
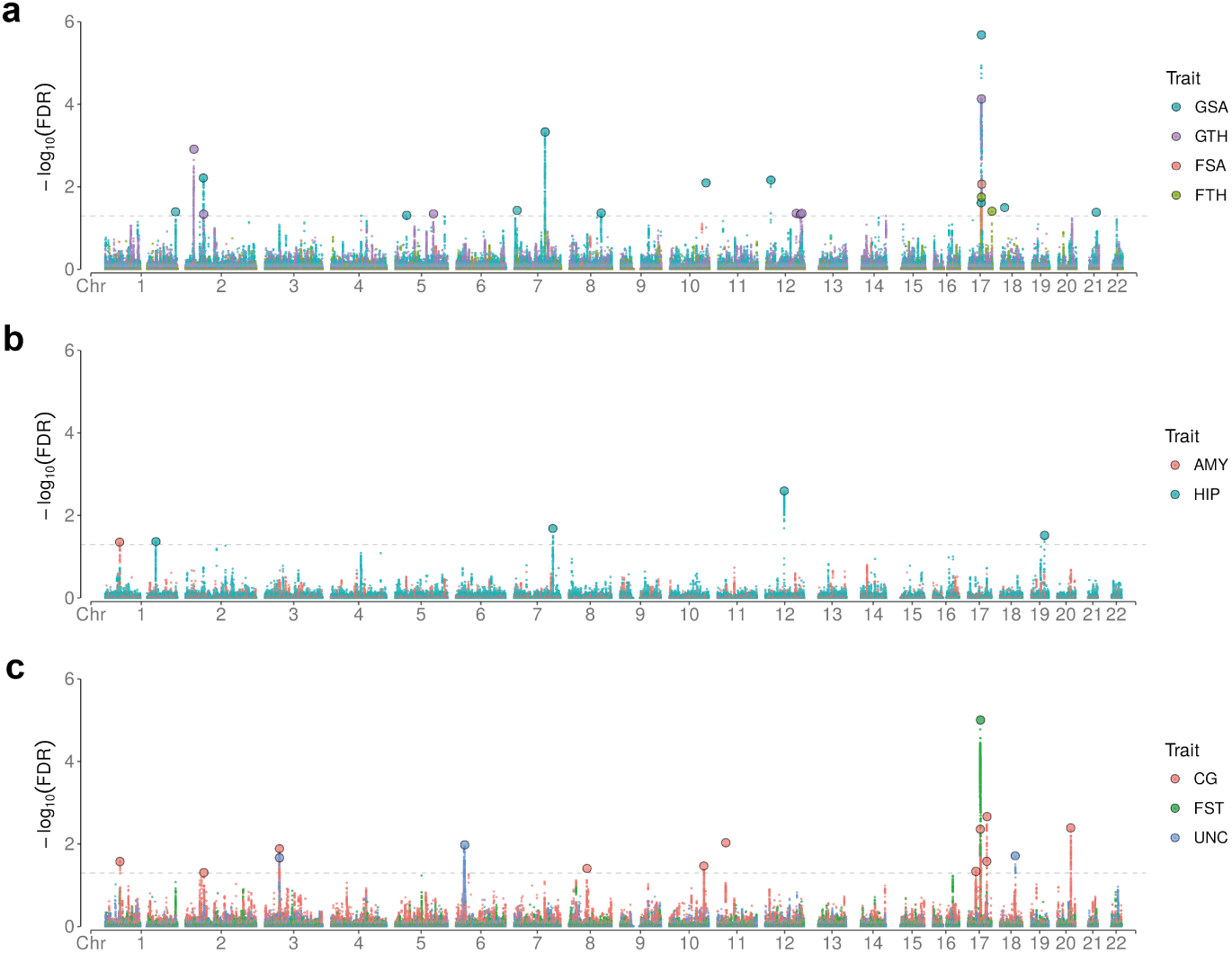
Chromosomal distribution of genetic loci jointly associated with PTSD and brain structure-related traits. The Manhattan plots show the common genetic variants jointly associated with PTSD and brain structure traits at conjFDR *<* 0.05. **a.** Results between PTSD and cortical traits. **b.**Results between PTSD and subcortical traits. **c.**Results between PTSD and white matter traits. Further details are provided in Table 2 and Supplementary Table 6-9. Abbreviations: GSA, global cortical surface area; GTH, global cortical thickness; FSA, frontal poles surface area; FTH, frontal poles thickness; AMY: amygdala; HIP: hippocampus; CG: cingulum bundle; FST: fornix/stria terminalis; UNC: uncinate fasciculus.

**Table 2.**
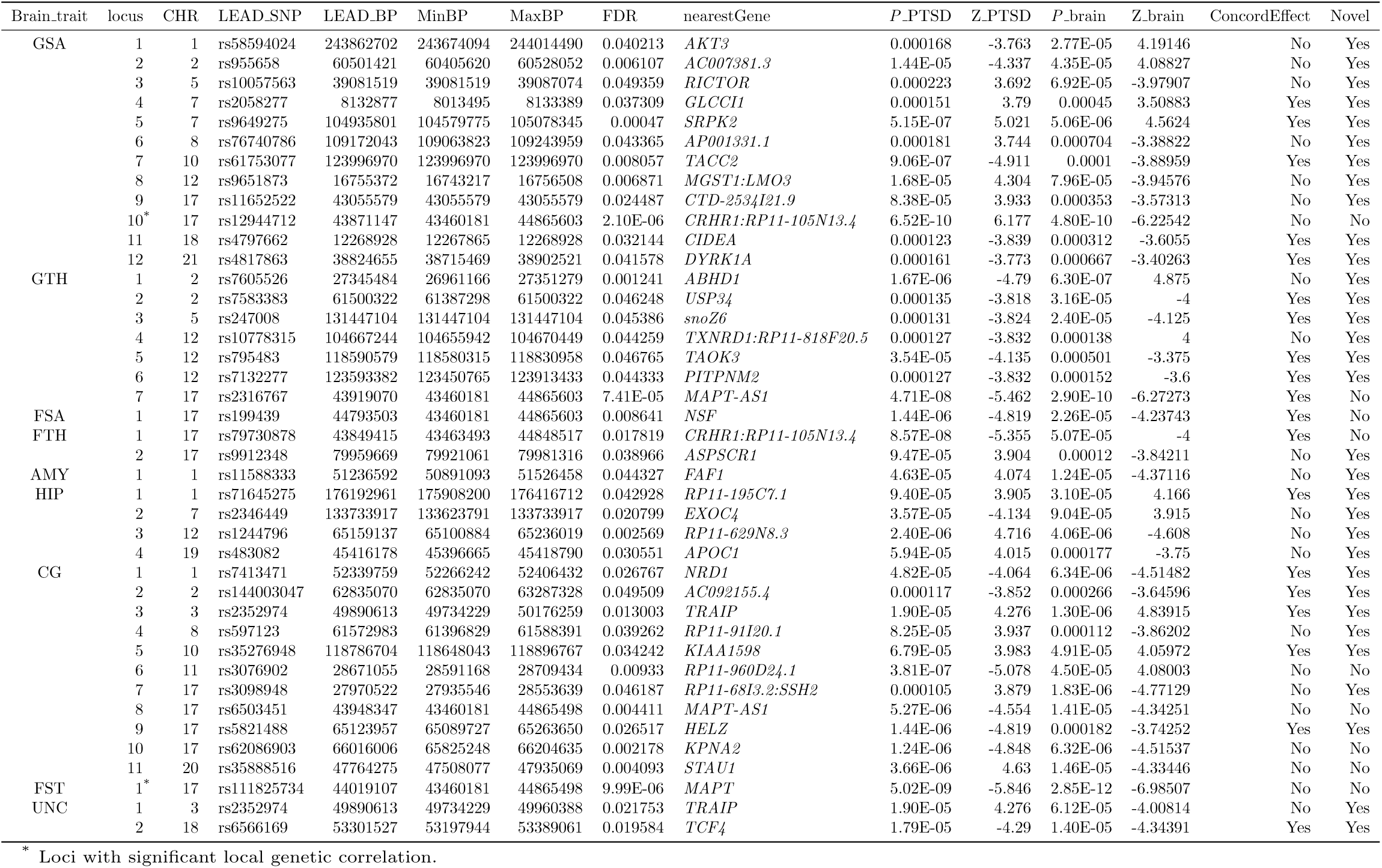
Lead genomic loci significantly associated with PTSD and brain structural traits at a conjFDR *<* 0.05.

### 3.5 Functional annotation and gene-set analysis

Functional annotation for all shared leader SNPs identified in conjFDR between PTSD and brain structure analysis showed that 76% were intergenic or intronic, while only 1 lead SNP was exonic and 8 SNPs were associated with nocoding-RNA. Four lead SNPs (rs955658, rs9651873, rs199439, and rs7413471) had a CADD score above the threshold score of 12.37, indicative of deleteriousness. Two of these were associated with GSA and showed opposite effects in PTSD and GSA, which was consistent with the conclusions of basic research. Moreover, 5 lead SNPs displayed “1f” in the RDB score, which means they were likely to affect binding and be linked to the expression of a gene target. Functional annotation for all SNPs in the loci shared between PTSD and brain structure (conjFDR *<* 0.05) displayed that 60.2% of them were intronic or intergenic (1692 of 2810), 1.4% were exonic (40 of 2810), and 33.9% were associated with nocoding-RNA (953 of 2810).

We applied FUMA to link the candidate SNPs (*r*^2^ *≥* 0.6) of 40 shared lead loci to 1,426 genes. It is worth noting that genes MAPT-AS1, SPPL2C, and CRHR1 could be annotated in all three ways and were related to five or more structure traits. These three genes were all located on chromosome 17. (Supplementary Table 7-10) Gene-set analysis of Gene Ontology (GO) for genes indicated by all SNPs in the loci shared between PTSD and brain structure, respectively, showed significantly associated 6 biological processes, 4 cellular components, and 8 molecular functions. (Supplementary Table 11)

### 3.6 Positive causal relationship from PTSD to FSA

We used bidirectional Mendelian randomization to explore whether the genetic overlap between two phenotypes suggests a causal relationship. Among all phenotype pairs, we only found a significant and consistent causal relationship from PTSD to FSA using 3 of 5 MR methods. (Fig 5). No obvious heterogeneity was detected in genetic variants associated with PTSD and FSA (Cochran’s Q = 10.18 and *P* = 0.60). The IVW method showed that genetically predicted higher PTSD risk was related to FSA (*β* 13.01; 95% CI, 4.49 to 21.53; *P* = 0.003). MR-RAPs and weighted median both demonstrated the effect of PTSD on FSA and provided evidence of the stability of the results of the IVW method. The results among other traits are detailed in Supplementary Table 12.

**Fig. 5.**
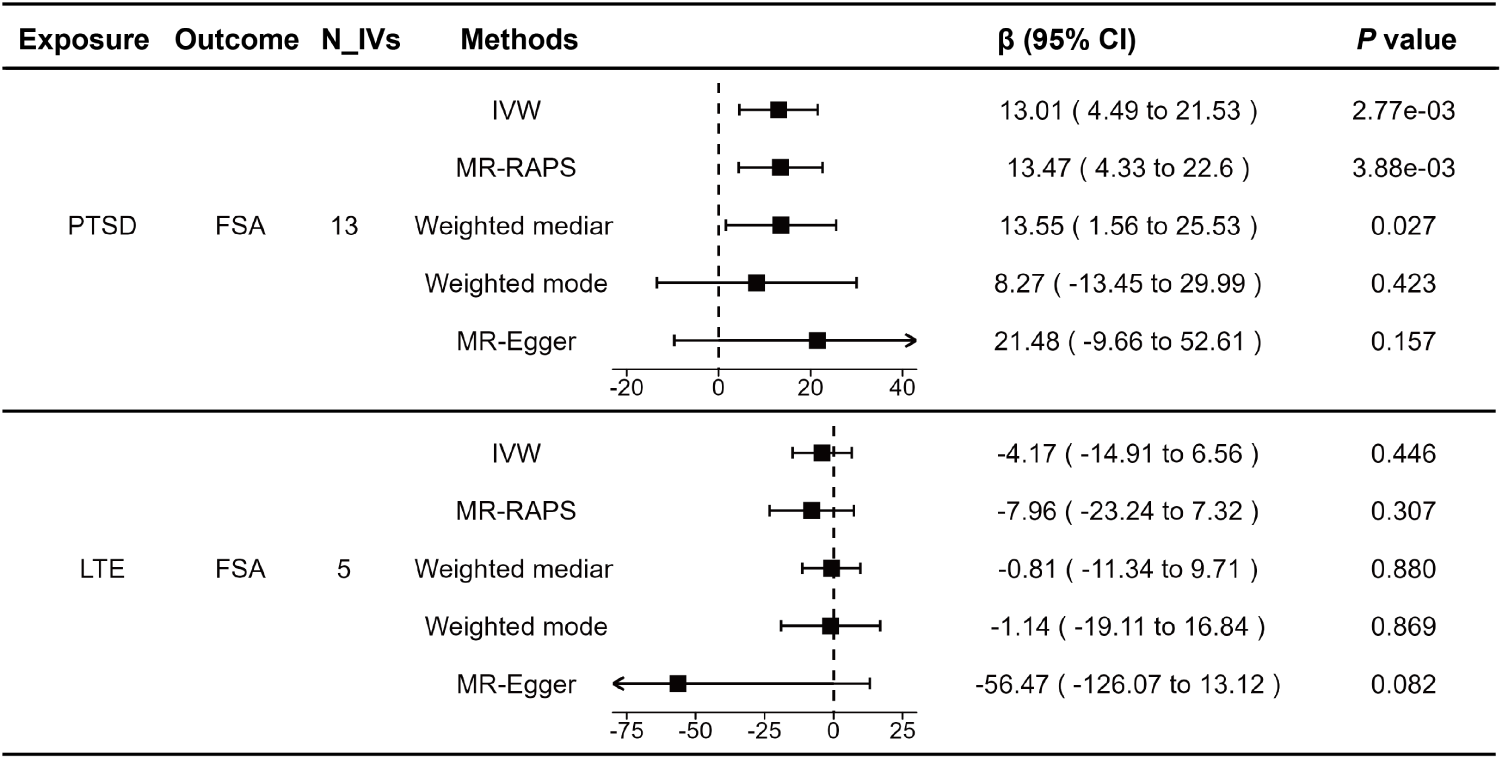
MR estimates of PTSD and LTE on FSA. Data are presented as β and 95% CI. N IVs: the number of instrument variants. Abbreviations: PTSD, post-traumatic stress disorder; LTE, lifetime trauma events; FSA, frontal poles surface area.

The scatter plots of SNP potential effects on PTSD versus FSA were demonstrated in Supplementary Fig. 4, with the slope of each representing the evaluated effect size per method. Among the 13 SNPs, only rs641325 was related to increased risks of PTSD and FSA. The result of the LOO analysis was presented in Supplementary Fig. 5, where no single SNP was driving the whole effect.

## 4 Discussion

In this study, we first conducted a meta-analysis on three accessible large-scale GWAS datasets, revealing 10 novel loci that were not identified in any of the three individual studies to enhance the statistical power. Then we utilized a series of genetically informed analyses to investigate the genetic association between PTSD and the brain structures of trauma-related neural circuits, using LTE as control. Our findings revealed varying degrees of relationship between PTSD and brain structures related to trauma circuits using different methods. These results provide new genetic evidence for PTSD and a deeper perspective into the pathogenesis.

Previous findings in animal studies and neuroimaging research regarding neural circuitry have been similarly corroborated on a genetic level. We have discovered genetic correlations or overlaps between PTSD and these brain regions at various levels. Additionally, previous studies have shown that there is a strong correlation between PTSD and LTE in both twin studies and genetic correlation studies.[44, 77] For most traits about mental health, *r_g_* with PTSD was also quite similar to *r_g_* with LTE[44]. In our analysis, there were more significant or stronger results for PTSD than LTE, indicating that trauma-related neural circuits were more related to the onset of PTSD than to the mediating effect of PTSD caused by being related to LTE. This further underscores the significance of neural circuits in PTSD.

We found that the heritability of SNPs associated with PTSD was significantly enriched in GTEx v8 “Brain Anterior cingulate cortex (BA24)”, “Brain Frontal Cortex (BA9)” and “Brain Cortex”, while there was no obvious founding in the SNPs of LTE. BA 24 is the ventral part of the brain anterior cingulate, which is connected with the amygdala and hippocampus, and it is involved in emotional tasks such as assessing the salience of emotion and motivational information.[78] BA9 is a cellularly defined part of the frontal cortex, and it is involved in short-term memory, suppressing sadness and recall which may be associated with the onset of PTSD.[79–81] These regions are anatomically close to the mPFC and its connections to the subcortex. The brain cortex covers a wider scope, involving attention, perception, consciousness, thinking, memory, and many other aspects. This suggests that the association of PTSD with its neural circuitry can indeed be explained at the genetic level. This provided a reference for our subsequent research on the cortex.

We studied the global and local genetic correlation between trauma-related traits and brain structure-related traits and found a weak global correlation between PTSD and GSA, although the results failed to pass FDR. Furthermore, we thought this might be due to mixed effect directions between the two traits. This was also confirmed in the subsequent LAVA local correlations and polygenic overlap results. Apart from FTH, we found 33 significant local bivariate correlations in 8 other pairs about PTSD. LAVA analysis showed a balanced mixture of concordant and disconcordant results throughout the genome. This suggested that different loci might be involved in different processes between disease and neurological traits, leading to different directions of influence. It was worth noting that the region in chr7:155280611-156344386 or chr17:43460501-4486583 was significant in both pairs about PTSD; the latter also coincided with the conjFDR results. Genes in these loci might play an important role in the genetic association between PTSD and brain structure traits.

We found that the trauma-related traits were considerably more polygenic (7.8K– 10K) than brain structure (0.3K–3.1K), reflecting that the mechanism of genetic factors in PTSD and LTE was far more complex than the mechanism in brain structure. LDSC was also used in MiXeR to estimate global correlation. In the previous part, when calculating global correlations, we used SNPs in HAPMAP3, and there was no obvious difference from the results calculated here. The results of bivariate MiXeR revealed that the genetic overlap between PTSD and brain structure accounts for at least 77%, some of these results were further supported by AIC values. Combining with the mixing effects we found in the conjFDR results, we inferred that there was genetic overlap without correlation[40] for PTSD and brain structure traits.

Through novelty search and functional annotation of 40 PTSD risk loci identified by conjFDR, we studied the possible mechanisms of PTSD and structural changes in neural circuits. 3p21.31 (lead SNP rs2352974) was a novel locus and was associated in both CG and UNC with PTSD. It was mapped to *TRAIP* positionally and to RBM6 and RNF123 by eQTL in the brain. The *TRAIP* gene is linked to the processes of DNA damage and repair.[82, 83] Specifically, *TRAIP* functions in the disentanglement of stalled replication forks during mitosis, thereby mitigating the occurrence of DNA bridges and premature loss of neural stem cells[84], so the damage to white matter in PTSD patients might be related to their neuron count. RBM6 and *TRAIP* have similar functions and are involved in the repair of DNA damage.[85] *RNF123* is linked to the ubiquitin-proteasome system and plays a role in depression.[86]. This process was confirmed in the gene-set analysis related to UNC, indicating that it was likely to be involved in the process of white matter damage in PTSD patients.

After annotating candidate genes from conjFDR results onto the genome using three methods, three genes (*MAPT-AS1*, *SPPL2C*, and *CRHR1*) were identified across 5 or more pairs, all of which are located on chromosome 17. Moreover, in this area, the conjFDR results of PTSD-GSA and PTSD-FST were consistent with the local correlation results of LAVA. *MAPT* is the gene responsible for encoding tau protein, which plays an important role in neurodegenerative diseases such as AD. The primary deposition sites for tau protein are the hippocampus and frontal lobe, precisely aligning with neural circuits implicated in trauma-related processes.[87, 88] *SPPL2C* is in the region of *MAPT*, is associated with *MAPT* expression in astrocytes.[89]. It has been observed that *CRHR1* antagonists can inhibit the activation of the CRH/NF-κB/BDNF pathway, thereby effectively preventing the rapid loss of synapses and memory impairment associated with trauma-induced delirium-like syndrome.[90] This may provide a new hypothesis for the occurrence of PTSD.

However, our research still has some limitations. First, there may be some trauma subjects might also be included in the neuroimaging samples, but we were unable to ascertain the influence of such overlap on our findings. Second, compared to schizophrenia[91] and bipolar disorder[92], PTSD exhibits lower heritability[93], so we have identified fewer loci associated with each phenotype in our study. And the summary we used exclusively focused on individuals of European ancestry. This limited scope may not fully capture the entirety of the genetic etiology involved. Future investigations may necessitate larger GWAS cohorts, encompassing diverse population groups, to corroborate our findings comprehensively. Then, the traits we selected mainly focused on the structure of the neural circuits rather than function. Some studies believe that PTSD is a disease caused by abnormal brain neural circuits.[15] Although there are GWAS on functional imaging, the phenotypic selection in these studies was derived through independent component analysis (ICA).[94] Consequently, the integration of phenotypes with neural circuits is challenging due to this chosen methodology. Last, we were unable to ascertain the causal relationship between trauma-related traits and brain structural traits, indicating the need for further research in this regard.

## 5 Conclusion

In this study, we have presented evidence of polygenic overlap between PTSD and trauma-related neural circuits. Forty overlapping loci and 1,426 genes were identified among nine phenotypes associated with brain structure in the context of PTSD. These findings contribute valuable insights into the shared genetic architecture between PTSD and its neural circuits, suggesting a common neurobiological foundation.

## Data Availability

All data produced in the present work are contained in the manuscript.

## Supplementary information

Supplementary data to this article can be found at Supplementary material and Supplementary tables.

## Acknowledgements

We express our gratitude to Professor Adam X. Maihofer for providing the GWAS summary data on PTSD and LTE. We also express our gratitude to the ENIGMA and Professor Bingxin Zhao et al. for their foundational research in the brain structural GWAS. The numerical calculations in this paper have been done on the supercomputing system in the Supercomputing Center of Wuhan University.

## Declarations

- Funding: This work was supported by grants from the National Natural Science Foundation of China (U21A20364) and the National Key R&D Program of China (2018YFC1314600).
- Conflict of interest: All authors declare that they have no competing interests.
- Ethics approval and consent to participate: All GWAS data sets included in this study were approved by the relevant ethics committees, and informed consent was obtained from all participants.
- Consent for publication: Not applicable
- Data availability: The PTSD GWAS data, encompassing the queues of both the PGC and the UK Biobank, were obtained through direct correspondence with the original authors.[44], and PTSD GWAS data from MVP and FinnGen can be respectively accessed at https://www.research.va.gov/mvp/(dbGaP Study Accession phs001672) and https://www.finngen.fi/en. The cortical and subcortical GWAS data were obtained from https://enigma.ini.usc.edu/, and the white matter GWAS data was from https://zenodo.org/records/4549730.
- Materials availability: Not applicable
- Code availability: All software used to conduct the analyses in this paper are freely available online: METAL: https://github.com/statgen/METAL. LDSC and LDSC-SEG: https://github.com/bulik/ldsc, LAVA: https://github.com/josefin-werme/LAVA, MiXeR: https://github.com/precimed/mixer, pleiofdr: https://github.com/precimed/pleiofdr, FUMA: https://fuma.ctglab.nl/.
- Author contribution: **Qian Gong** (Data curation, Formal analysis, Investigation, Methodology, Visualization, Writing original draft). **Honggang Lv** (Data curation, Formal analysis, Methodology, Editing). **Lijun Kang** (Investigation, Editing). **Simeng Ma** (Data curation, Investigation). **Nan Zhang** (Investigation). **Xinhui Xie** (Investigation, Editing). **Enqi Zhou** (Investigation). **Zipeng Deng** (Investigation). **Jiewei Liu** (Data curation, Formal analysis, Methodology, Editing). **Zhongchun Liu** (Methodology, Investigation, Resources, Supervision, Editing).

## Notes

### Competing Interest Statement

The authors have declared no competing interest.

### Funding Statement

This study was funded by the National Natural Science Foundation of China (U21A20364) and the National Key R&D Program of China (2018YFC1314600)

## References

[1] Pai, A., Suris, A. M. & North, C. S. Posttraumatic Stress Disorder in the DSM-5: Controversy, Change, and Conceptual Considerations. Behav Sci (Basel) 7, 7 (2017).

[2] Shalev, A., Liberzon, I. & Marmar, C. Post-Traumatic Stress Disorder. N Engl J Med 376, 2459–2469 (2017).

[3] McLaughlin, K. A. et al. Subthreshold posttraumatic stress disorder in the world health organization world mental health surveys. Biol Psychiatry 77, 375–384 (2015).

[4] Kessler, R. C. & Wang, P. S. The descriptive epidemiology of commonly occurring mental disorders in the United States. Annu Rev Public Health 29, 115–129 (2008).

[5] Kilpatrick, D. G. et al. National estimates of exposure to traumatic events and PTSD prevalence using DSM-IV and DSM-5 criteria. J Trauma Stress 26, 537– 547 (2013).

[6] Maddox, S. A., Hartmann, J., Ross, R. A. & Ressler, K. J. Deconstructing the Gestalt: Mechanisms of Fear, Threat, and Trauma Memory Encoding. Neuron 102, 60–74 (2019).

[7] Milad, M. R. et al. Neurobiological basis of failure to recall extinction memory in posttraumatic stress disorder. Biol Psychiatry 66, 1075–1082 (2009).

[8] Iqbal, J., Huang, G.-D., Xue, Y.-X., Yang, M. & Jia, X.-J. The neural circuits and molecular mechanisms underlying fear dysregulation in posttraumatic stress disorder. Front Neurosci 17, 1281401 (2023).

[9] Rauch, S. L., Shin, L. M. & Phelps, E. A. Neurocircuitry models of posttraumatic stress disorder and extinction: Human neuroimaging research–past, present, and future. Biol Psychiatry 60, 376–382 (2006).

[10] Harnett, N. G., Goodman, A. M. & Knight, D. C. PTSD-related neuroimaging abnormalities in brain function, structure, and biochemistry. Exp Neurol 330, 113331 (2020).

[11] Krabbe, S., Gründemann, J. & Lüthi, A. Amygdala Inhibitory Circuits Regulate Associative Fear Conditioning. Biol Psychiatry 83, 800–809 (2018).

[12] Tovote, P., Fadok, J. P. & Lüthi, A. Neuronal circuits for fear and anxiety. Nat Rev Neurosci 16, 317–331 (2015).

[13] Asede, D., Bosch, D., Lüthi, A., Ferraguti, F. & Ehrlich, I. Sensory inputs to intercalated cells provide fear-learning modulated inhibition to the basolateral amygdala. Neuron 86, 541–554 (2015).

[14] Dejean, C. et al. Neuronal Circuits for Fear Expression and Recovery: Recent Advances and Potential Therapeutic Strategies. Biol Psychiatry 78, 298–306 (2015).

[15] Patel, R., Spreng, R. N., Shin, L. M. & Girard, T. A. Neurocircuitry models of posttraumatic stress disorder and beyond: A meta-analysis of functional neuroimaging studies. Neurosci Biobehav Rev 36, 2130–2142 (2012).

[16] Hayes, J. P., Hayes, S. M. & Mikedis, A. M. Quantitative meta-analysis of neural activity in posttraumatic stress disorder. Biol Mood Anxiety Disord 2, 9 (2012).

[17] Qin, C. et al. Dorsal Hippocampus to Infralimbic Cortex Circuit is Essential for the Recall of Extinction Memory. Cereb Cortex 31, 1707–1718 (2021).

[18] Cleren, C. et al. Low-frequency stimulation of the ventral hippocampus facilitates extinction of contextual fear. Neurobiol Learn Mem 101, 39–45 (2013).

[19] Guan, Y., Chen, X., Zhao, B., Shi, Y. & Han, F. What Happened in the Hippocampal Axon in a Rat Model of Posttraumatic Stress Disorder. Cell Mol Neurobiol 42, 723–737 (2022).

[20] Hayes, J. P. et al. Reduced hippocampal and amygdala activity predicts memory distortions for trauma reminders in combat-related PTSD. J Psychiatr Res 45, 660–669 (2011).

[21] Carríon, V. G., Haas, B. W., Garrett, A., Song, S. & Reiss, A. L. Reduced hippocampal activity in youth with posttraumatic stress symptoms: An FMRI study. J Pediatr Psychol 35, 559–569 (2010).

[22] Do-Monte, F. H., Quiñones-Laracuente, K. & Quirk, G. J. A temporal shift in the circuits mediating retrieval of fear memory. Nature 519, 460–463 (2015).

[23] Do-Monte, F. H., Manzano-Nieves, G., Quiñones-Laracuente, K., Ramos-Medina, L. & Quirk, G. J. Revisiting the role of infralimbic cortex in fear extinction with optogenetics. J Neurosci 35, 3607–3615 (2015).

[24] Garfinkel, S. N. et al. Impaired contextual modulation of memories in PTSD: An fMRI and psychophysiological study of extinction retention and fear renewal. J Neurosci 34, 13435–13443 (2014).

[25] Aupperle, R. L. et al. Dorsolateral prefrontal cortex activation during emotional anticipation and neuropsychological performance in posttraumatic stress disorder. Arch Gen Psychiatry 69, 360–371 (2012).

[26] Rougemont-Bücking, A., et al. Altered processing of contextual information during fear extinction in PTSD: An fMRI study. CNS Neurosci Ther 17, 227–236 (2011).

[27] Nicholson, A. A. et al. The Dissociative Subtype of Posttraumatic Stress Disorder: Unique Resting-State Functional Connectivity of Basolateral and Centromedial Amygdala Complexes. Neuropsychopharmacology 40, 2317–2326 (2015).

[28] Gilmartin, M. R., Balderston, N. L. & Helmstetter, F. J. Prefrontal cortical regulation of fear learning. Trends Neurosci 37, 455–464 (2014).

[29] Harnett, N. G., Ference, E. W., Knight, A. J. & Knight, D. C. White matter microstructure varies with post-traumatic stress severity following medical trauma. Brain Imaging Behav 14, 1012–1024 (2020).

[30] Koch, S. B. J. et al. Decreased uncinate fasciculus tract integrity in male and female patients with PTSD: A diffusion tensor imaging study. J Psychiatry Neurosci 42, 331–342 (2017).

[31] Molńar, Z., et al. New insights into the development of the human cerebral cortex. J Anat 235, 432–451 (2019).

[32] Batouli, S. A. H., Trollor, J. N., Wen, W. & Sachdev, P. S. The heritability of volumes of brain structures and its relationship to age: A review of twin and family studies. Ageing Res Rev 13, 1–9 (2014).

[33] Batouli, S. A. H. et al. Heritability of brain volumes in older adults: The Older Australian Twins Study. Neurobiol Aging 35, 937.e5–18 (2014).

[34] Kanchibhotla, S. C. et al. Genetics of ageing-related changes in brain white matter integrity - a review. Ageing Res Rev 12, 391–401 (2013).

[35] Grasby, K. L. et al. The genetic architecture of the human cerebral cortex. Science 367, eaay6690 (2020).

[36] Satizabal, C. L. et al. Genetic architecture of subcortical brain structures in 38,851 individuals. Nat Genet 51, 1624–1636 (2019).

[37] Hibar, D. P. et al. Novel genetic loci associated with hippocampal volume. Nat Commun 8, 13624 (2017).

[38] Zhao, B. et al. Common genetic variation influencing human white matter microstructure. Science 372, eabf3736 (2021).

[39] Stauffer, E.-M. et al. The genetic relationships between brain structure and schizophrenia. Nat Commun 14, 7820 (2023).

[40] Cheng, W. et al. Shared genetic architecture between schizophrenia and subcortical brain volumes implicates early neurodevelopmental processes and brain development in childhood. Mol Psychiatry 27, 5167–5176 (2022).

[41] Wu, B.-S. et al. Genome-wide association study of cerebellar white matter microstructure and genetic overlap with common brain disorders. Neuroimage 269, 119928 (2023).

[42] Shang, M.-Y. et al. Genetic associations between bipolar disorder and brain structural phenotypes. Cereb Cortex bhad014 (2023).

[43] Ressler, K. J. et al. Post-traumatic stress disorder: Clinical and translational neuroscience from cells to circuits. Nat Rev Neurol 18, 273–288 (2022).

[44] Maihofer, A. X. et al. Enhancing Discovery of Genetic Variants for Posttraumatic Stress Disorder Through Integration of Quantitative Phenotypes and Trauma Exposure Information. Biol Psychiatry 91, 626–636 (2022).

[45] Nievergelt, C. M. et al. International meta-analysis of PTSD genome-wide association studies identifies sex- and ancestry-specific genetic risk loci. Nat Commun 10, 4558 (2019).

[46] Frei, O. et al. Bivariate causal mixture model quantifies polygenic overlap between complex traits beyond genetic correlation. Nat Commun 10, 2417 (2019).

[47] Smeland, O. B. et al. Discovery of shared genomic loci using the conditional false discovery rate approach. Hum Genet 139, 85–94 (2020).

[48] Watanabe, K., Taskesen, E., van Bochoven, A. & Posthuma, D. Functional mapping and annotation of genetic associations with FUMA. Nat Commun 8, 1826 (2017).

[49] Stein, M. B. et al. Genome-wide association analyses of post-traumatic stress disorder and its symptom subdomains in the Million Veteran Program. Nat Genet 53, 174–184 (2021).

[50] Harrington, K. M. et al. Validation of an Electronic Medical Record-Based Algorithm for Identifying Posttraumatic Stress Disorder in U.S. Veterans. J Trauma Stress 32, 226–237 (2019).

[51] Willer, C. J., Li, Y. & Abecasis, G. R. METAL: Fast and efficient meta-analysis of genomewide association scans. Bioinformatics 26, 2190–2191 (2010).

[52] Desikan, R. S. et al. An automated labeling system for subdividing the human cerebral cortex on MRI scans into gyral based regions of interest. Neuroimage 31, 968–980 (2006).

[53] Rakic, P. Specification of cerebral cortical areas. Science 241, 170–176 (1988).

[54] Brodmann, K. Vergleichende lokalisationslehre der grosshirnrinde. Leipzig: Barth JA (1905).

[55] Patenaude, B., Smith, S. M., Kennedy, D. N. & Jenkinson, M. A Bayesian model of shape and appearance for subcortical brain segmentation. Neuroimage 56, 907–922 (2011).

[56] Fischl, B. et al. Whole brain segmentation: Automated labeling of neuroanatomical structures in the human brain. Neuron 33, 341–355 (2002).

[57] Grieve, S. M., Williams, L. M., Paul, R. H., Clark, C. R. & Gordon, E. Cognitive aging, executive function, and fractional anisotropy: A diffusion tensor MR imaging study. AJNR Am J Neuroradiol 28, 226–235 (2007).

[58] Finucane, H. K. et al. Heritability enrichment of specifically expressed genes identifies disease-relevant tissues and cell types. Nat Genet 50, 621–629 (2018).

[59] GTEx Consortium. The GTEx Consortium atlas of genetic regulatory effects across human tissues. Science 369, 1318–1330 (2020).

[60] Bulik-Sullivan, B. et al. An atlas of genetic correlations across human diseases and traits. Nat Genet 47, 1236–1241 (2015).

[61] 1000 Genomes Project Consortium et al. A global reference for human genetic variation. Nature 526, 68–74 (2015).

[62] Werme, J., van der Sluis, S., Posthuma, D. & de Leeuw, C. A. An integrated framework for local genetic correlation analysis. Nat Genet 54, 274–282 (2022).

[63] Andreassen, O. A. et al. Improved detection of common variants associated with schizophrenia and bipolar disorder using pleiotropy-informed conditional false discovery rate. PLoS Genet 9, e1003455 (2013).

[64] Buniello, A. et al. The NHGRI-EBI GWAS Catalog of published genome-wide association studies, targeted arrays and summary statistics 2019. Nucleic Acids Res 47, D1005–D1012 (2019).

[65] Rentzsch, P., Witten, D., Cooper, G. M., Shendure, J. & Kircher, M. CADD: Predicting the deleteriousness of variants throughout the human genome. Nucleic Acids Res 47, D886–D894 (2019).

[66] Boyle, A. P. et al. Annotation of functional variation in personal genomes using RegulomeDB. Genome Res 22, 1790–1797 (2012).

[67] Roadmap Epigenomics Consortium et al. Integrative analysis of 111 reference human epigenomes. Nature 518, 317–330 (2015).

[68] GTEx Consortium et al. Genetic effects on gene expression across human tissues. Nature 550, 204–213 (2017).

[69] Wang, D. et al. Comprehensive functional genomic resource and integrative model for the human brain. Science 362, eaat8464 (2018).

[70] Burgess, S., Thompson, S. G. & CRP CHD Genetics Collaboration. Avoiding bias from weak instruments in Mendelian randomization studies. Int J Epidemiol 40, 755–764 (2011).

[71] Kamat, M. A. et al. PhenoScanner V2: An expanded tool for searching human genotype-phenotype associations. Bioinformatics 35, 4851–4853 (2019).

[72] Myers, T. A., Chanock, S. J. & Machiela, M. J. LDlinkR: An R Package for Rapidly Calculating Linkage Disequilibrium Statistics in Diverse Populations. Front Genet 11, 157 (2020).

[73] Burgess, S. et al. Using published data in Mendelian randomization: A blueprint for efficient identification of causal risk factors. Eur J Epidemiol 30, 543–552 (2015).

[74] Bowden, J., Davey Smith, G., Haycock, P. C. & Burgess, S. Consistent Estimation in Mendelian Randomization with Some Invalid Instruments Using a Weighted Median Estimator. Genet Epidemiol 40, 304–314 (2016).

[75] Bowden, J., Davey Smith, G. & Burgess, S. Mendelian randomization with invalid instruments: Effect estimation and bias detection through Egger regression. Int J Epidemiol 44, 512–525 (2015).

[76] Verbanck, M., Chen, C.-Y., Neale, B. & Do, R. Detection of widespread horizontal pleiotropy in causal relationships inferred from Mendelian randomization between complex traits and diseases. Nat Genet 50, 693–698 (2018).

[77] Stein, M. B., Jang, K. L., Taylor, S., Vernon, P. A. & Livesley, W. J. Genetic and environmental influences on trauma exposure and posttraumatic stress disorder symptoms: A twin study. Am J Psychiatry 159, 1675–1681 (2002).

[78] Bush, G., Luu, P. & Posner, M. I. Cognitive and emotional influences in anterior cingulate cortex. Trends Cogn Sci 4, 215–222 (2000).

[79] Babiloni, C. et al. Human cortical responses during one-bit delayed-response tasks: An fMRI study. Brain Res Bull 65, 383–390 (2005).

[80] Kaur, S. et al. Cingulate cortex anatomical abnormalities in children and adolescents with bipolar disorder. Am J Psychiatry 162, 1637–1643 (2005).

[81] Tulving, E. et al. Neuroanatomical correlates of retrieval in episodic memory: Auditory sentence recognition. Proc Natl Acad Sci U S A 91, 2012–2015 (1994).

[82] Wu, R. A. et al. TRAIP is a master regulator of DNA interstrand crosslink repair. Nature 567, 267–272 (2019).

[83] Wu, R. A., Pellman, D. S. & Walter, J. C. The Ubiquitin Ligase TRAIP: Double-Edged Sword at the Replisome. Trends Cell Biol 31, 75–85 (2021).

[84] O’Neill, R. S. & Rusan, N. M. Traip controls mushroom body size by suppressing mitotic defects. Development 149, dev199987 (2022).

[85] Awwad, S. W., Darawshe, M. M., Machour, F. E., Arman, I. & Ayoub, N. Recruitment of RBM6 to DNA Double-Strand Breaks Fosters Homologous Recombination Repair. Mol Cell Biol 43, 130–142 (2023).

[86] Teyssier, J.-R., Rey, R., Ragot, S., Chauvet-Gelinier, J.-C. & Bonin, B. Correlative gene expression pattern linking RNF123 to cellular stress-senescence genes in patients with depressive disorder: Implication of DRD1 in the cerebral cortex. J Affect Disord 151, 432–438 (2013).

[87] Strang, K. H., Golde, T. E. & Giasson, B. I. MAPT mutations, tauopathy, and mechanisms of neurodegeneration. Lab Invest 99, 912–928 (2019).

[88] Leveille, E., Ross, O. A. & Gan-Or, Z. Tau and MAPT genetics in tauopathies and synucleinopathies. Parkinsonism Relat Disord 90, 142–154 (2021).

[89] He, L. et al. Exome-wide age-of-onset analysis reveals exonic variants in ERN1 and SPPL2C associated with Alzheimer’s disease. Transl Psychiatry 11, 146 (2021).

[90] Cursano, S. et al. A CRHR1 antagonist prevents synaptic loss and memory deficits in a trauma-induced delirium-like syndrome. Mol Psychiatry 26, 3778– 3794 (2021).

[91] Trubetskoy, V. et al. Mapping genomic loci implicates genes and synaptic biology in schizophrenia. Nature 604, 502–508 (2022).

[92] Stahl, E. A. et al. Genome-wide association study identifies 30 loci associated with bipolar disorder. Nat Genet 51, 793–803 (2019).

[93] Nievergelt, C. M. et al. Discovery of 95 PTSD loci provides insight into genetic architecture and neurobiology of trauma and stress-related disorders. medRxiv 2023.08.31.23294915 (2023).

[94] Zhao, B. et al. Common variants contribute to intrinsic human brain functional networks. Nat Genet 54, 508–517 (2022).

