## Supplementary Fig for "The genetic relationships between post-traumatic stress disorder and its corresponding neural circuit structures"

**This PDF file includes:**

Supplementary Notes

Figs. S1 to S5

**Other Supplementary Materials for this manuscript include the following:**

**Supplementarytable.xlsx:** Tables. S1 to S12

Supplementary Notes

**GWAS summary details**

**PTSD.** The first GWAS summary mentioned in the original test was from the meta-analysis across 51 cohorts of European ancestry, including the Psychiatric Genomics Consortium (PGC, freeze2 PGC-PTSD meta-analysis), and the UK Biobank, encompassing a total of 182,199 samples[1]. Nineteen methods were used to evaluate PTSD, the most common being the Clinician-Administered PTSD Scale and PTSD Checklist, and 91% of participants with PTSD were using the PTSD symptom score, while the others used the case/control status[1]. Specific methods of participant and trait selection can be found in the original article[1,2]. The second summary was from MVP[3]. PTSD in MVP was a diagnosis of case/control applying the algorithm based on the electronic medical record. There were 36,301 cases and 178,107 controls. The third summary was from FinnGen, which was also defined as a case/control status, with 2,282 cases, 337,577 controls.

**LTE.** The LTE summary was from UK Biobank with 132,988 samples[1]. The researchers constructed a count measure of LTE from 8 trauma items of the self-reported retrospective trauma screener from the UKBB mental-health questionnaire.

**Cortical.** These GWAS summaries were from ENIGMA3 (https://enigma.ini.usc.edu/). It comprised results from 33,992 participants of European ancestry (23,909 from 49 cohorts in the ENIGMA consortium and 10,083 from the UK Biobank)[4]. The traits of this study involve the surface area (SA) and thickness (TH) of 34 brain regions of the Desikan-Killiany (DK) atlas. These brain regions were divided according to the gyral-based neuroanatomical regions[5].

**Subcortical.** The GWAS for the volume of amygdala was from a formal study including 53 cohorts from the CHARGE consortium, ENIGMA consortium, and UK Biobank[6]. The traits were defined as the mean volume (in cm^3^) of the left and right amygdala[6]. Segmentation of brain regions was done according to the freely available and in-house segmentation methods[6]. GWAS for the volume of hippocampus was obtained from a GWAS study of 33,536 individuals in the ENIGMA Consortium and the CHARGE Consortium[7]. Hippocampal volumes were also estimated using the automated segmentation algorithm in FMRIB Software Library (FSL) and FreeSurfer[8,9].

**White-matter** GWAS summary data for white-matter was obtained from a GWAS of dMRI data from 43,802 individuals across five data resources[10]. This article included 5 DTI indicators of 21 white matter pathways. We chose fractional anisotropy as the primary DTI indicator because it has received the most attention in brain white matter research.

References

1.	Maihofer AX, Choi KW, Coleman JRI, Daskalakis NP, Denckla CA, Ketema E, et al. Enhancing Discovery of Genetic Variants for Posttraumatic Stress Disorder Through Integration of Quantitative Phenotypes and Trauma Exposure Information. Biol Psychiatry 2022;91:626–36.

2.	Nievergelt CM, Maihofer AX, Klengel T, Atkinson EG, Chen C-Y, Choi KW, et al. International meta-analysis of PTSD genome-wide association studies identifies sex- and ancestry-specific genetic risk loci. Nat Commun 2019;10:4558.

3.	Harrington KM, Quaden R, Stein MB, Honerlaw JP, Cissell S, Pietrzak RH, et al. Validation of an Electronic Medical Record-Based Algorithm for Identifying Posttraumatic Stress Disorder in U.S. Veterans. J Trauma Stress 2019;32:226–37.

4.	Grasby KL, Jahanshad N, Painter JN, Colodro-Conde L, Bralten J, Hibar DP, et al. The genetic architecture of the human cerebral cortex. Science 2020;367:eaay6690.

5.	Desikan RS, Ségonne F, Fischl B, Quinn BT, Dickerson BC, Blacker D, et al. An automated labeling system for subdividing the human cerebral cortex on MRI scans into gyral based regions of interest. Neuroimage 2006;31:968–80.

6.	Satizabal CL, Adams HHH, Hibar DP, White CC, Knol MJ, Stein JL, et al. Genetic architecture of subcortical brain structures in 38,851 individuals. Nat Genet 2019;51:1624–36.

7.	Hibar DP, Adams HHH, Jahanshad N, Chauhan G, Stein JL, Hofer E, et al. Novel genetic loci associated with hippocampal volume. Nat Commun 2017;8:13624.

8.	Patenaude B, Smith SM, Kennedy DN, Jenkinson M. A Bayesian model of shape and appearance for subcortical brain segmentation. Neuroimage 2011;56:907–22.

9.	Fischl B, Salat DH, Busa E, Albert M, Dieterich M, Haselgrove C, et al. Whole brain segmentation: automated labeling of neuroanatomical structures in the human brain. Neuron 2002;33:341–55.

10.	Zhao B, Li T, Yang Y, Wang X, Luo T, Shan Y, et al. Common genetic variation influencing human white matter microstructure. Science 2021;372:eabf3736.

Fig. S1 Q-Q plot of the meta-analysis of PTSD. The red dashed line represents the predicted P-value, and the blue framed line shows the 95% confidence interval.

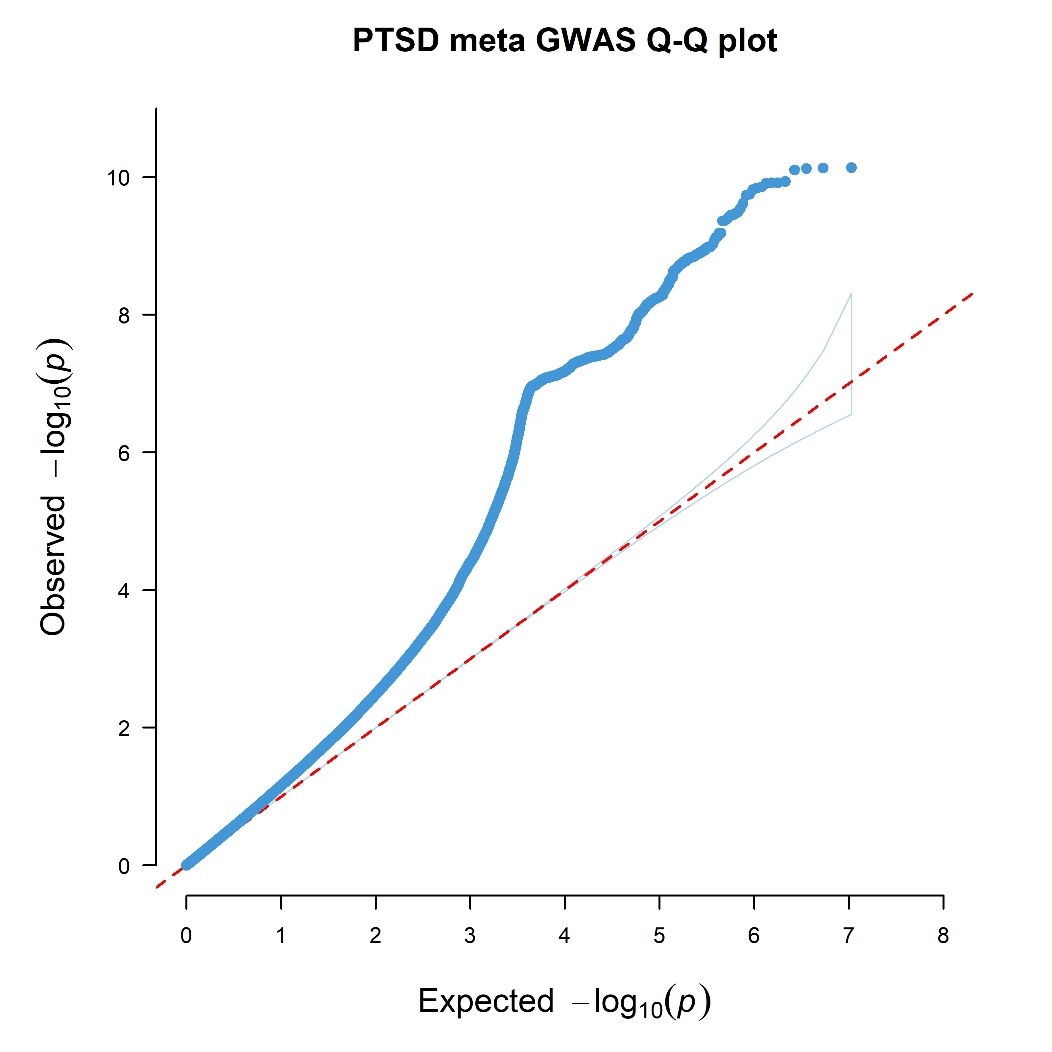

Fig. S2 The MiXeR-estimated heritability, polygenicity, and discoverability for each phenotype. The SNP heritability is a sum of effects across all trait-inﬂuencing variants. The polygenicity of each trait is represented by the number of trait-inﬂuencing variants to explain 90% heritability. The discoverability of each trait is deﬁned as effect size per trait-inﬂuencing variant. PTSD: post-traumatic stress disorder; LTE: lifetime trauma events; GSA: global surface area. GTH: global thickness; FSA: frontal pole surface area; FTH: frontal pole thickness; AMY: amygdala; HIP: Hippocampus; CG: cingulum; FST: fornix and stria terminalis; UNC: uncinate fasciculus.

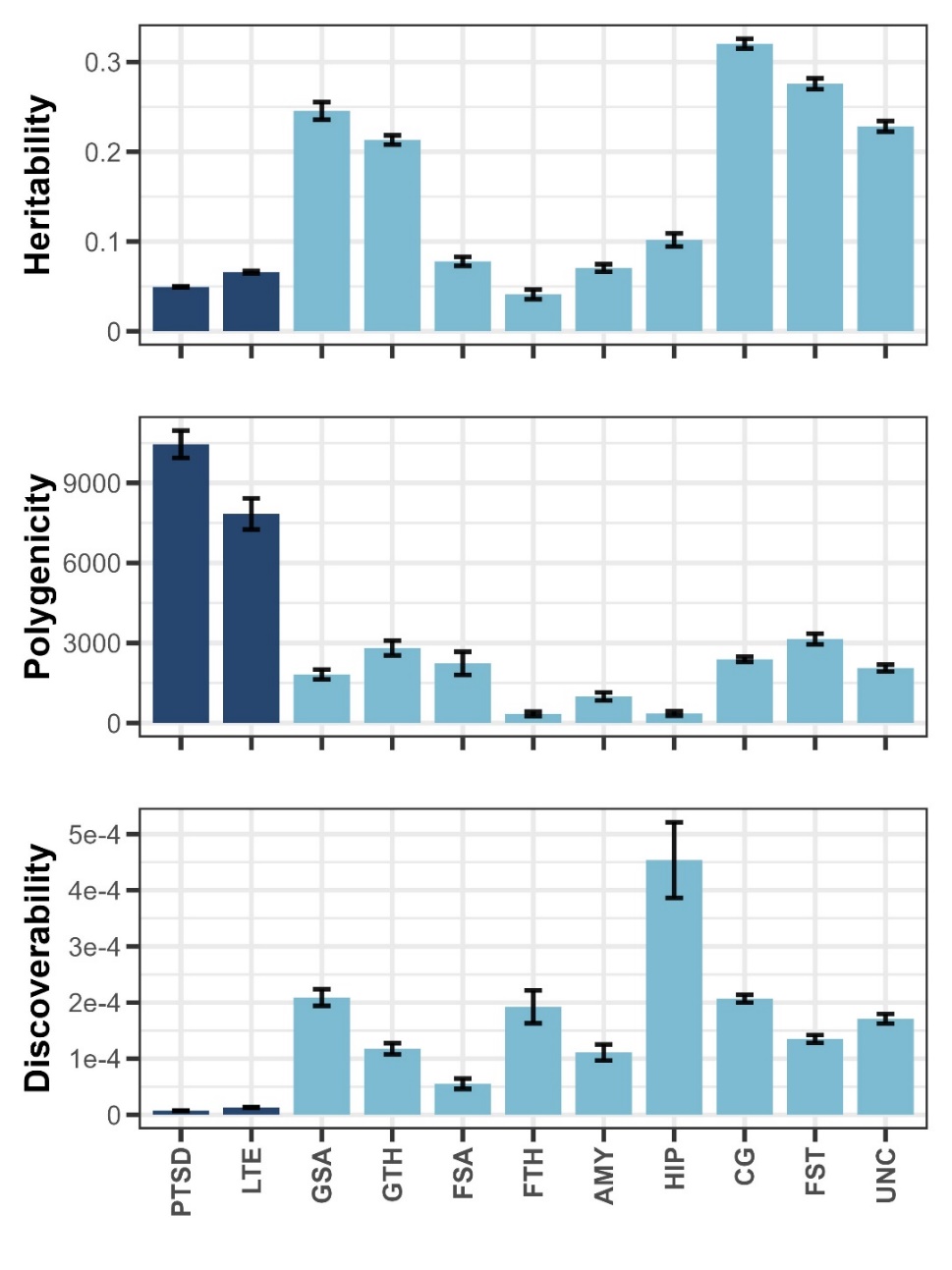

Fig. S3 The polygenic overlap between trauma-related traits and brain structure-related traits. For each pair of traits, MiXeR bivariate analysis generates: (1) a Venn plot to show the number of shared and trait-specific trait-influencing variants; (2) conditional QQ plots to reveal the cross-trait SNP enrichment; (3) negative log-likelihood plot to indicate the performance of the best model vs. min and max.

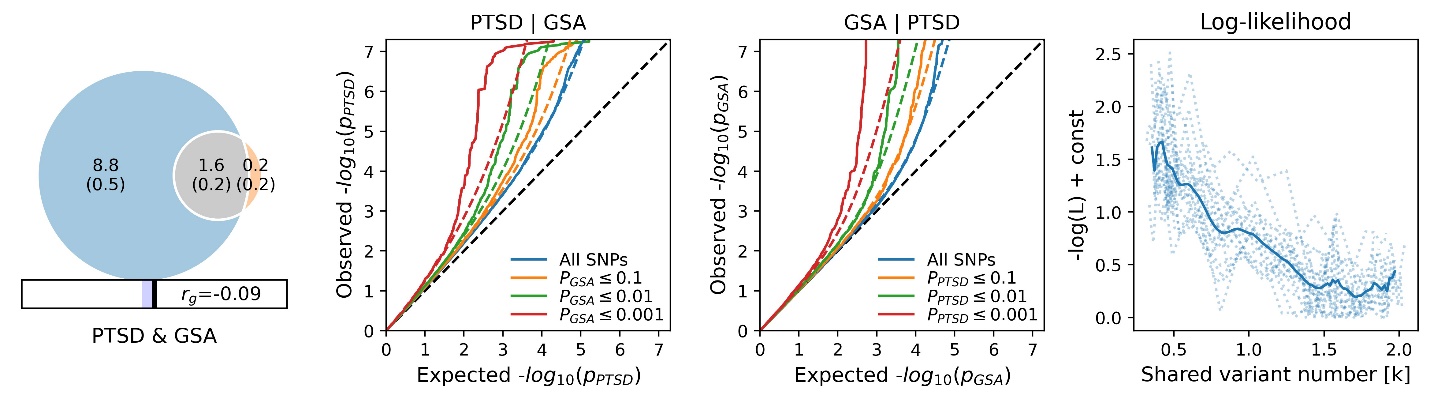

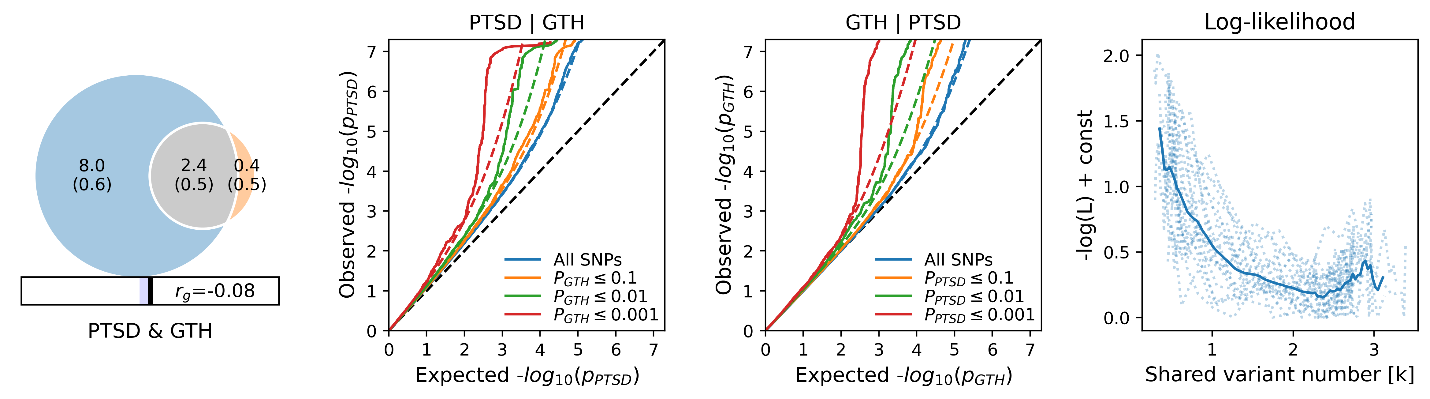

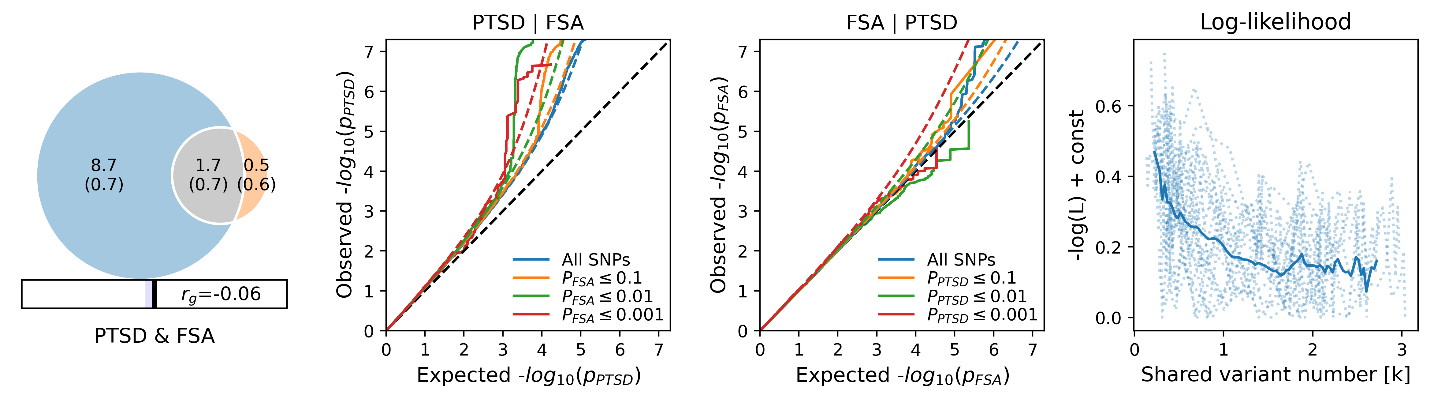

Continued on next page.

Continued from previous page.

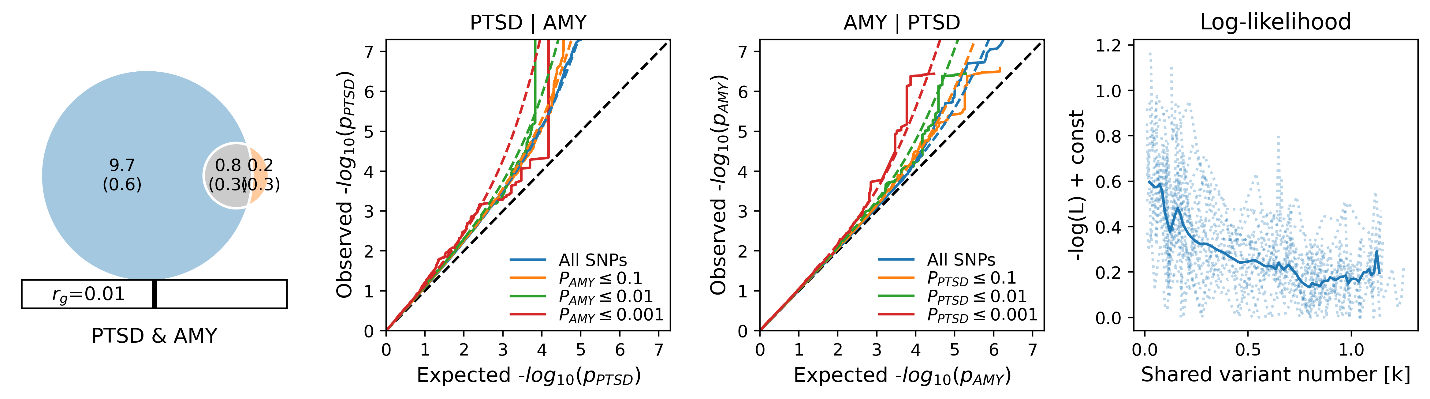

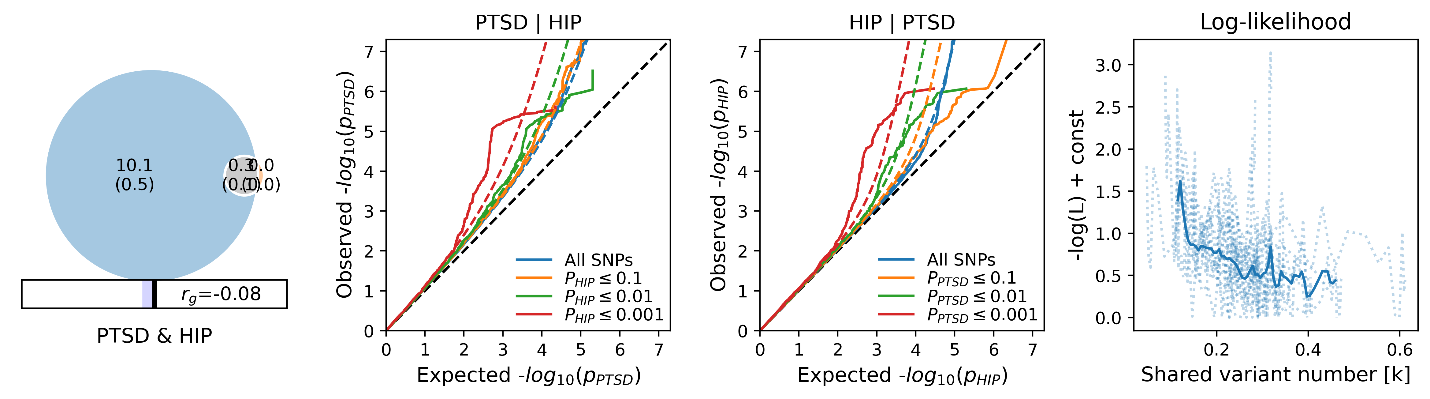

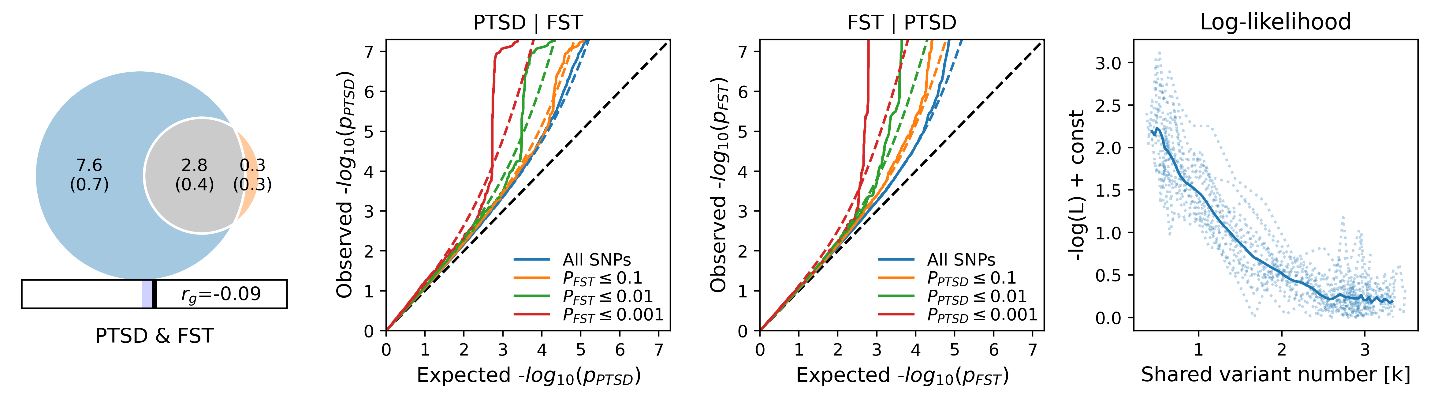

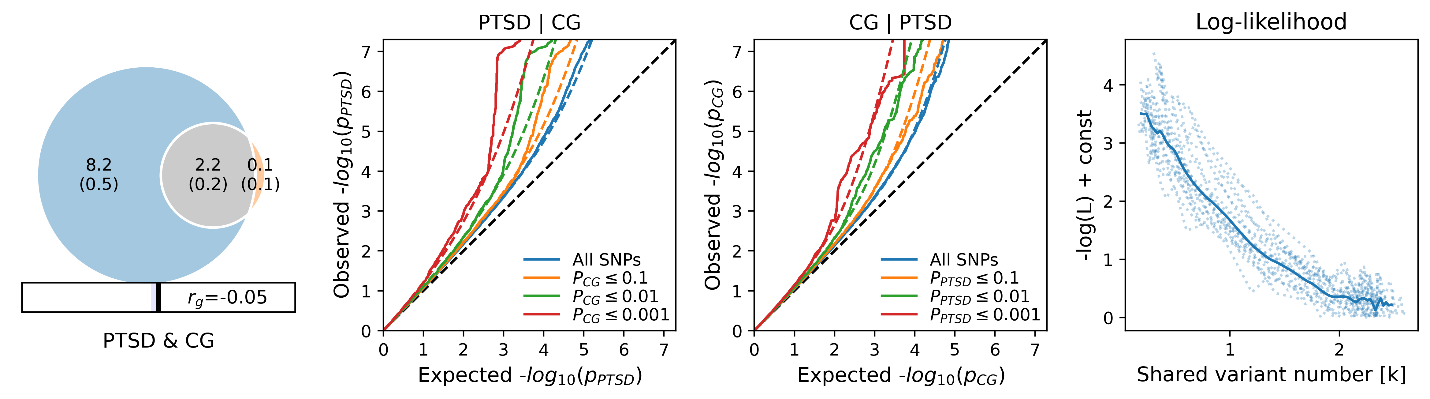

Continued on next page.

Continued from previous page.

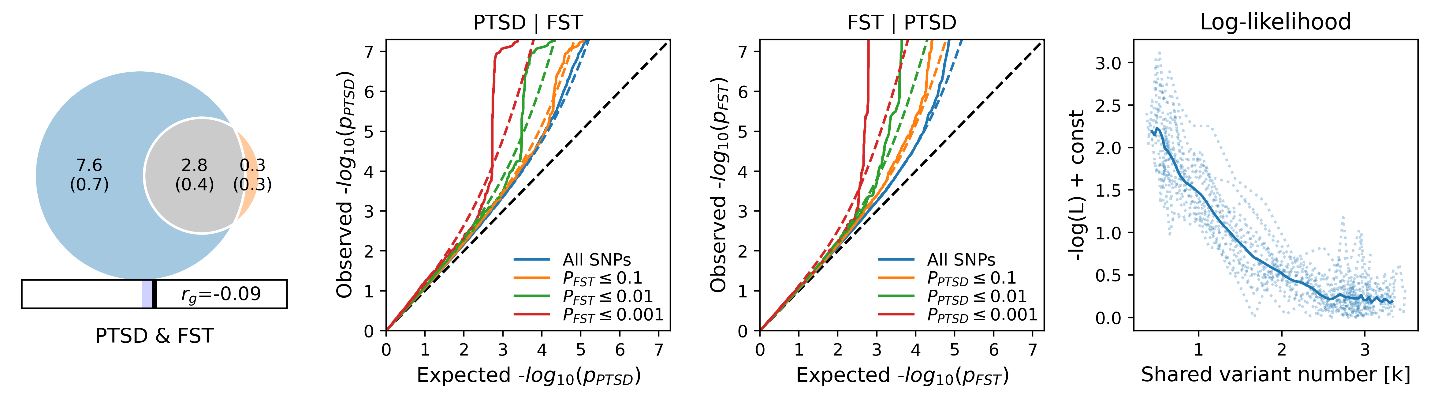

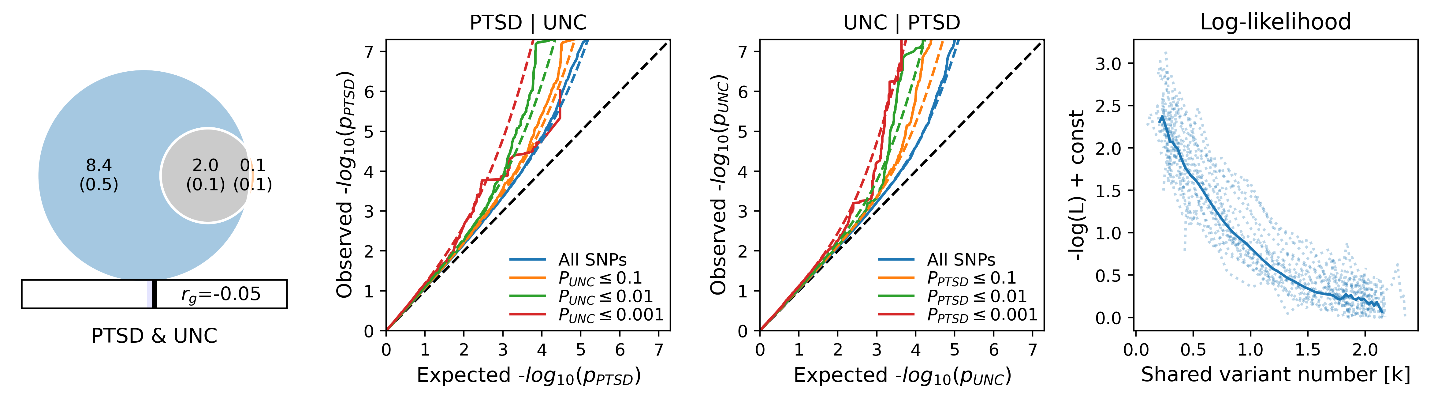

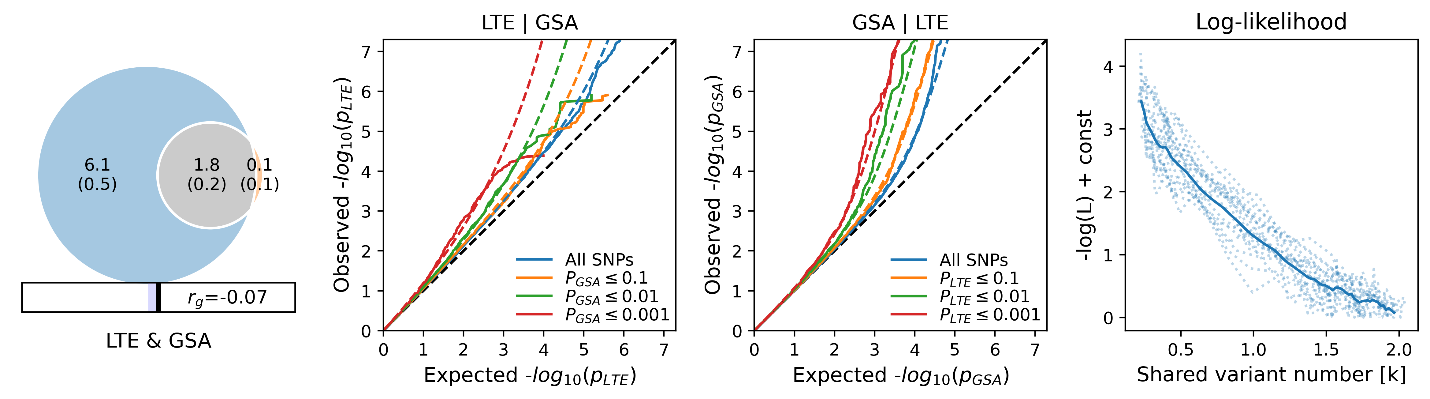

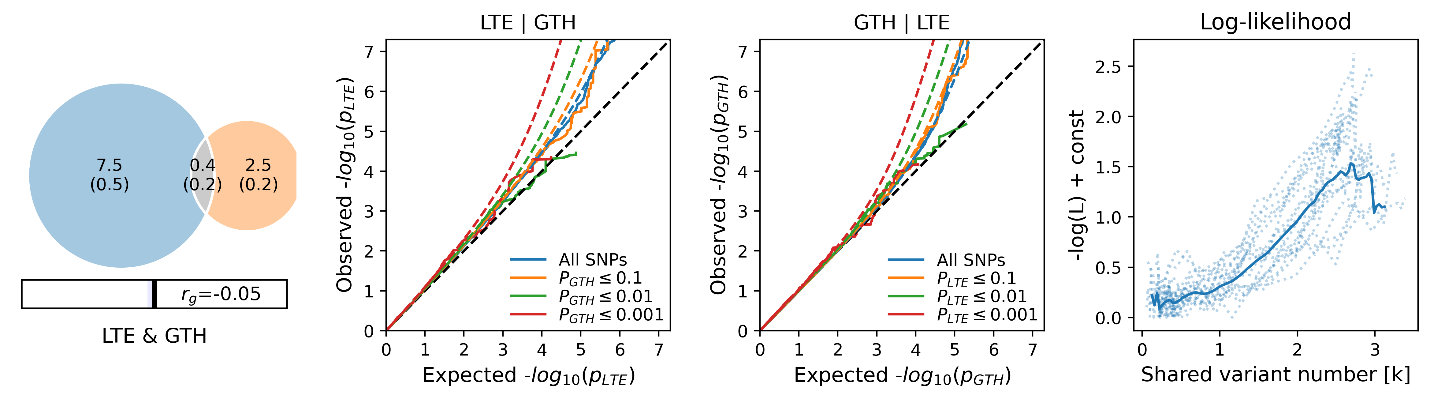

Continued on next page.

Continued from previous page.

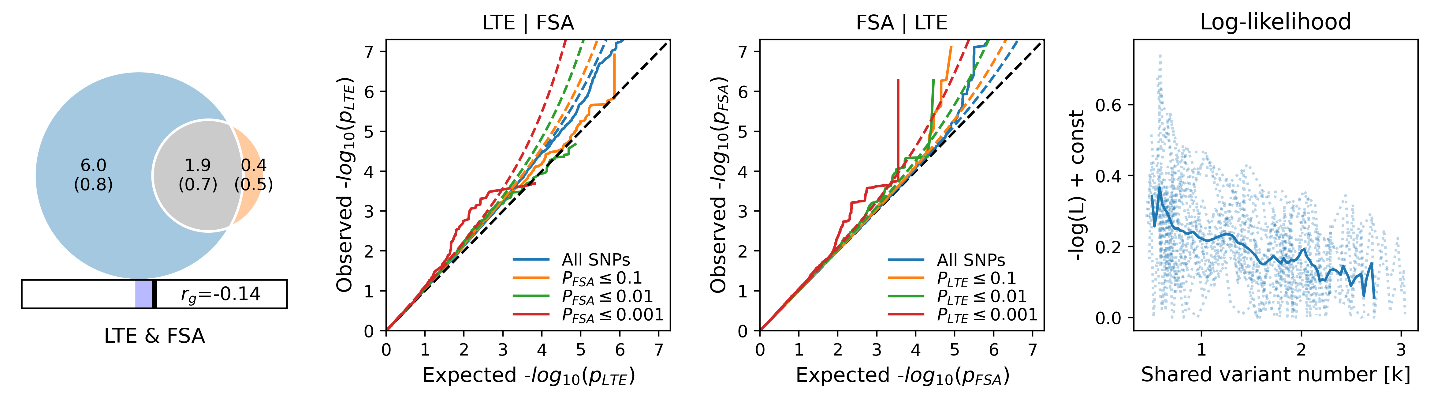

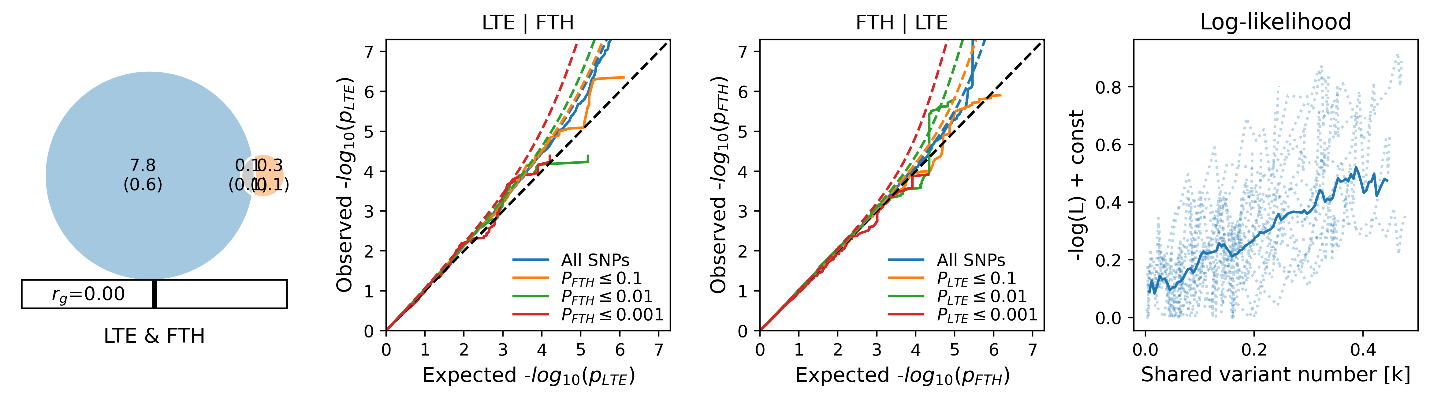

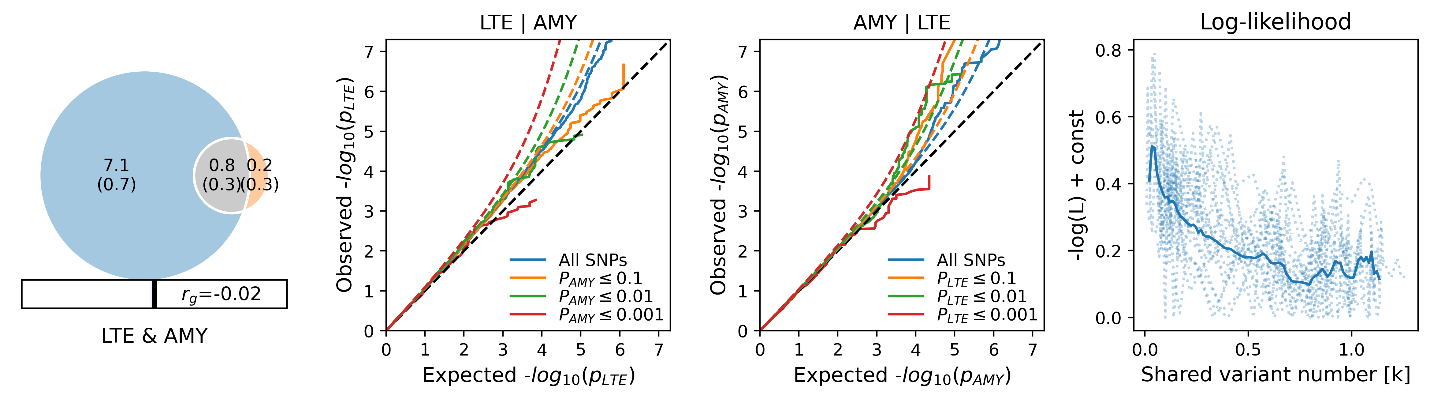

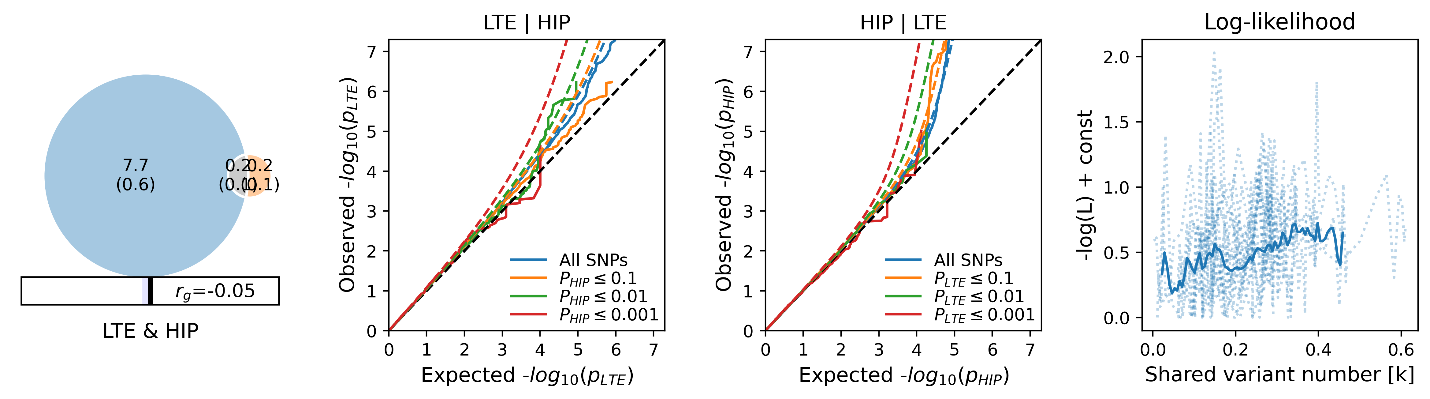

Continued on next page.

Continued from previous page.

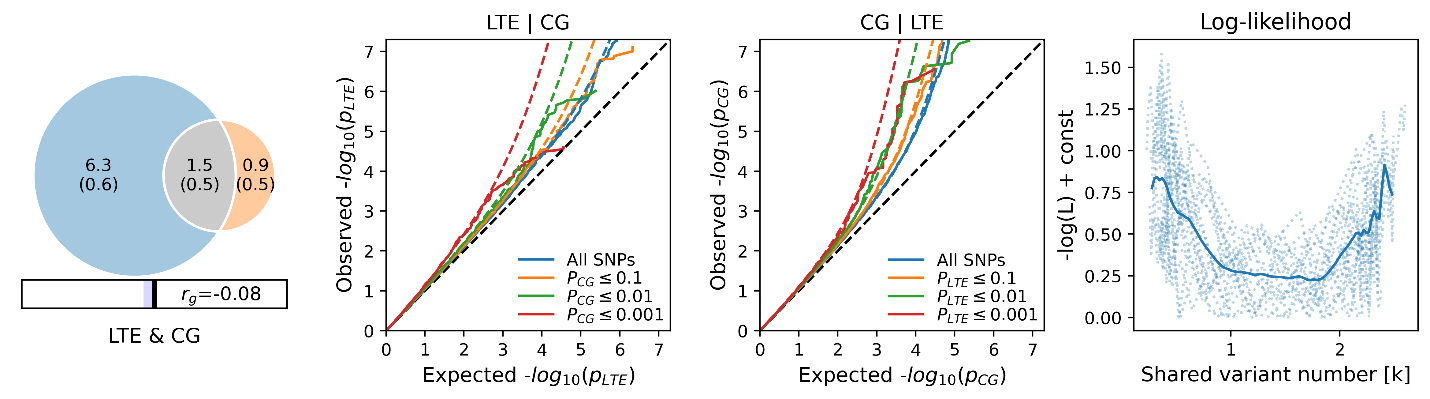

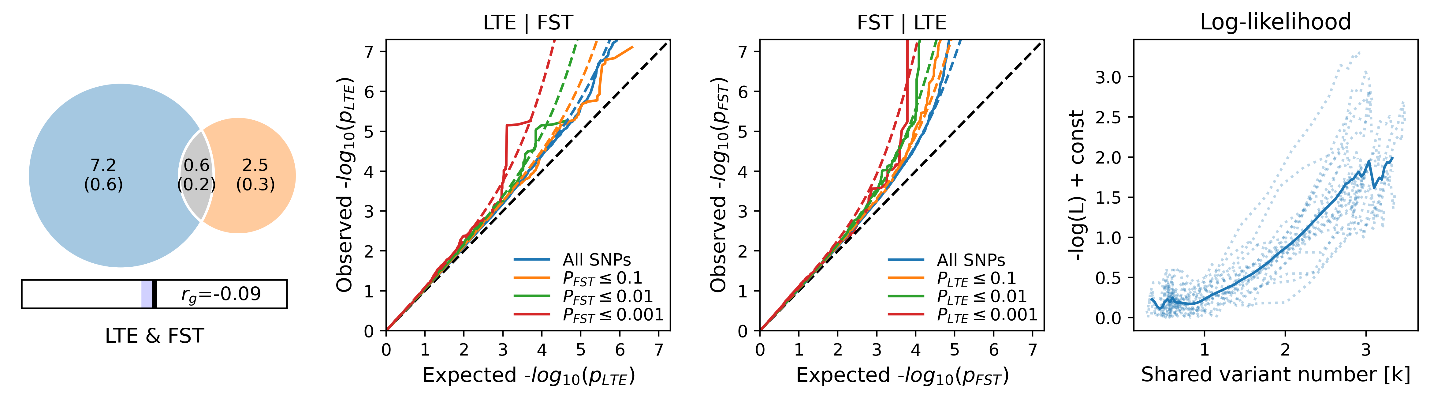

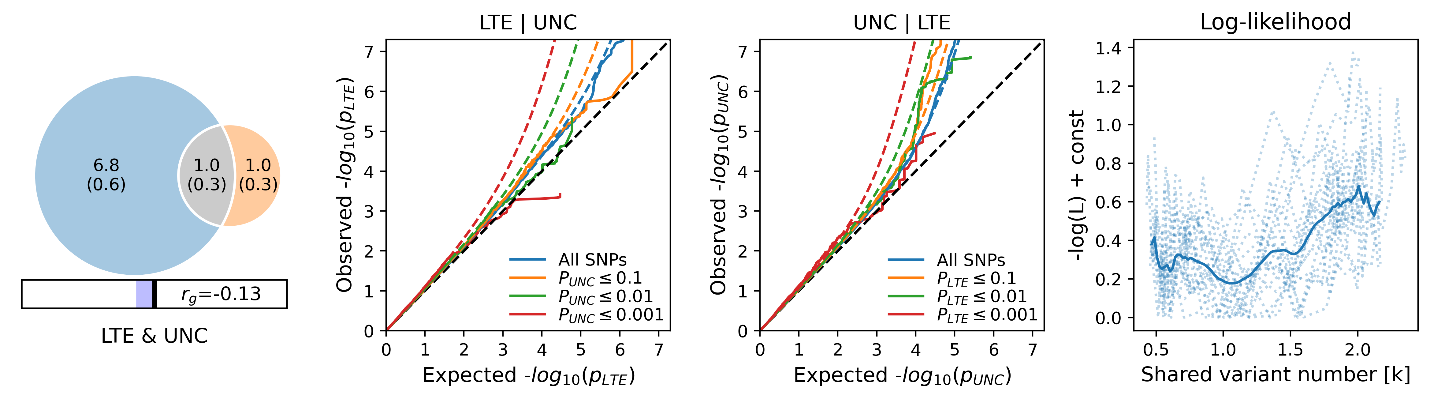

Fig. S4. Scatter plots of SNP effects on PTSD versus FSA. PTSD: post-traumatic stress disorder; FSA: frontal pole surface area.

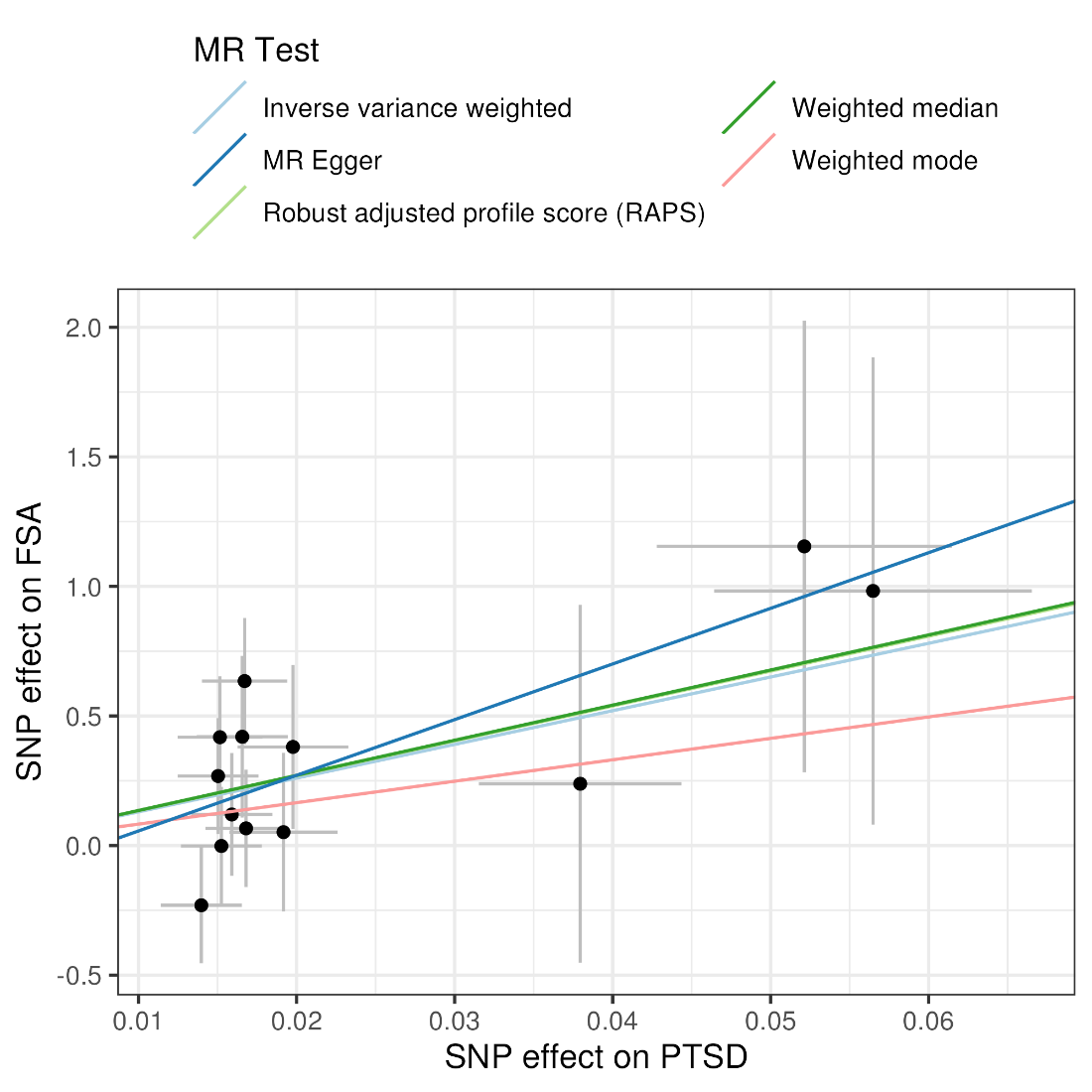

Fig. S5. The leave-one-out estimate of PTSD versus FSA. Data are presented as β with a 95% confidence interval. PTSD: post-traumatic stress disorder; FSA: frontal pole surface area.

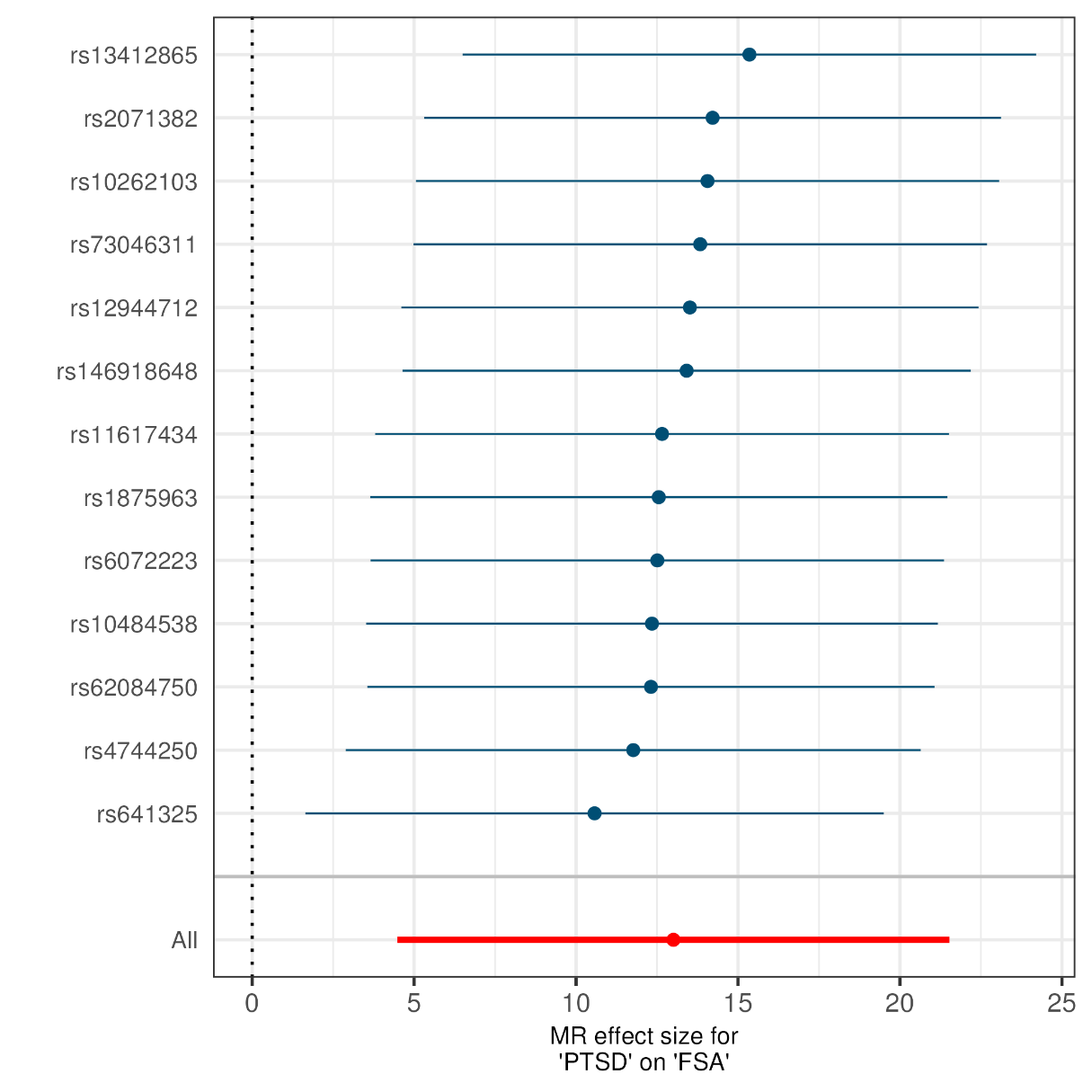

Supplementary tables (separate file)

| Table S1 | Results from tissue-type-specific enrichment of SNPs heritability using LDSC-SEG. |
| --- | --- |
| Table S2 | Results from global correlation using LDSC. |
| Table S3 | Results from local correlation using LAVA. |
| Table S4 | Results from univariate MiXeR. |
| Table S5 | Results from bivariate MiXeR. |
| Table S6 | Distinct genomic loci shared between trauma-related and brain structure-related traits at conjFDR<0.05. |
| Table S7 | All SNPs jointly associated with PTSD or LTE and cortical traits having a conjFDR <0.05 and an r2 > 0.6 with one of the independent significant SNPs. |
| Table S8 | All SNPs jointly associated with PTSD or LTE and subcortical traits having a conjFDR <0.05 and an r2 > 0.6 with one of the independent significant SNPs. |
| Table S9 | All SNPs jointly associated with PTSD or LTE and subcortical traits having a conjFDR <0.05 and an r2 > 0.6 with one of the independent significant SNPs. |
| Table S10 | Genes indicated by SNPs in the loci shared between PTSD and brain structure. |
| Table S11 | Gene Ontology (GO) for genes indicated by all SNPs in the loci shared between PTSD and brain structure. |
| Table S12 | Results from bidirectional two-sample Mendelian randomization analysis. |
